# Loss-of-function mutation in *IKZF2* leads to immunodeficiency with dysregulated germinal center reactions and reduction of MAIT cells

**DOI:** 10.1101/2021.08.25.21262015

**Authors:** Iivo Hetemäki, Meri Kaustio, Matias Kinnunen, Nelli Heikkilä, Salla Keskitalo, Simo Miettinen, Joona Sarkkinen, Virpi Glumoff, Noora Andersson, Kaisa Kettunen, Reetta Vanhanen, Katariina Nurmi, Kari K Eklund, Johannes Dunkel, Mikko Mäyränpää, Heinrich Schlums, T. Petteri Arstila, Kai Kisand, Yenan T. Bryceson, Pärt Peterson, Ulla Otava, Jaana Syrjänen, Janna Saarela, Markku Varjosalo, Eliisa Kekäläinen

## Abstract

The IKAROS family transcription factors regulate lymphocyte development. Loss-of-function variants in *IKZF1* cause primary immunodeficiency, but IKAROS family members *IKZF2* and *IKZF3* have not yet been associated with immunodeficiency yet. Here, we describe a pedigree with a heterozygous truncating variant in *IKZF2*, encoding the translational activator and repressor HELIOS which is highly expressed in regulatory T cells and effector T cells, particularly of the CD8^+^ T cell lineage. Protein-protein interaction analysis revealed that the variant abolished HELIOS dimerizations as well as binding to members of the Mi-2/NuRD chromatin remodeling complex. Patients carrying the *IKZF2* variant presented with a combined immunodeficiency phenotype characterized by recurrent upper respiratory infections, thrush and mucosal ulcers, as well as chronic lymphadenopathy. With extensive immunophenotyping, functional assays, and transcriptional analysis we show that reduced HELIOS expression was associated with chronic T cell activation and increased production of pro-inflammatory cytokines both in effector and regulatory T cells. Lymph node histology from patients indicated dysregulated germinal center reactions. Moreover, affected individuals displayed profoundly reduced circulating MAIT cell numbers. In summary, we show that this novel loss-of-function variant in HELIOS leads to an immunodeficiency with signs of immune overactivation.

**One sentence summary:** Truncating variant of HELIOS causes immunodeficiency with signs of immune overactivation.

## Introduction

The *IKZF2* gene encodes for the zinc-finger protein HELIOS that can act both as an activator and repressor of transcription. HELIOS is a member of the IKAROS family of transcription factors, which all share the same structure of two Krüppel-like zinc-finger domains. The N-terminal domain of the protein is required for DNA binding and the C-terminal domain mediates homo- as well as hetero-dimerization with other IKAROS family members, IKAROS and AIOLOS (*1*). IKAROS family members have a wide role on the development and function of the immune system. They appear to function through orchestrating chromatin remodeling, in which their interactions with the nucleosome remodeling and histone deacetylase (NuRD) complex, one of the major transcriptional corepressor complexes in mammalian cells, are essential (*2–4*). Both IKAROS and HELIOS are proto-oncogenes for hematological malignancies - IKAROS in acute lymphoblastic leukemia and HELIOS in T cell leukemias and acute myeloid leukemia (*4–6*). Heterozygous variants in the *IKZF1* gene causing haploinsufficiency of IKAROS have been recently shown to cause common variable immunodeficiency -like syndrome with variable clinical phenotypes (*7*), while dominant negative mutations are associated with T, B, and myeloid cell combined immunodeficiency (*8*).

Helios expression is mostly limited to the T cell lineage but young *Ikzf2* knockout mice show no clear immunological phenotype even though a large fraction of homozygous pups perishes for unknown reasons before weaning(*9*). Older *Ikzf2*^-/-^ mice develop an autoimmune phenotype characterized by autoantibodies and dysregulated germinal center reactions (*10, 11*). Helios is highly expressed in both murine and human T regulatory cells (Tregs) and therefore most studies have focused on its role in peripheral immune tolerance. Helios stabilizes the non-inflammatory phenotype of Tregs, possibly via STAT-5 mediated signaling and prevents IL-2 production in Tregs by epigenetic silencing (*12*). In selective knock-out models and human memory Tregs, Helios-negative Tregs produce more proinflammatory cytokines than their Helios-expressing counterparts (*13, 14*). However, the suppressive capacity of Tregs is not severely impaired in Helios^-/-^ mice (*11*).

In addition to the constitutively high expression of HELIOS in Tregs, the expression is also induced after TCR-mediated activation in both Tregs and effector T cells (*15, 16*). Other factors controlling the expression are unknown, but involvement of NFkappaB transcription factor has been suggested (*17*). Studies on T cell exhaustion with LCMV murine infection model identified Helios as one of the most important transcription factors differentiating exhausted virus specific T cells from naive and memory cells (*18, 19*). It is thus evident that HELIOS has a significant role in regulating effector T cell activity during immune responses.

Thus far no germline *IKZF2* variants have been described in humans with a primary immunodeficiency disease (PID). Here we describe a heterozygous *IKZF2* loss-of-function variant in a single family, causing an immunodeficiency with increased immune activation and profound reduction of Mucosal associated invariant T (MAIT) cells. Affected patients have lymphadenopathy with dysregulated germinal centers and aberrations in antibody production reminiscent of the Helios knock-out mouse phenotype. Our results emphasize the importance of the HELIOS’ protein binding domains in mediating its functions in both effector and regulatory T cells.

## Results

### A novel *IKZF2* variant associated with symptoms of immunodeficiency and immune dysregulation

The index patient, a female in her late 30’s, (patient 1) was referred to our immunodeficiency clinic due to chronic vulvovaginal *Candida albicans* infection, recurrent vulvar and oral mucosal aphtae, chronic lymphadenopathy, and recurrent upper respiratory infections (Table 1). In initial examinations she was diagnosed with hypogammaglobulinemia and is now receiving immunoglobulin replacement therapy. Her relative, a male in his 60’s, (patient 2) has suffered from recurrent pneumonias, lichen planus, and oral thrush. He was diagnosed with Hodgkin’s lymphoma at his 30s and also had chronic lymphadenopathy without lymphoma relapse. More detailed case reports are provided by the authors upon reasonable request.

**Table 1.** Clinical presentation and immunological phenotype of patients with the *IKZF2* p.Y200X variant

|  | Patient<br>1 | Patient<br>2 | Reference<br>values (unit) |
| --- | --- | --- | --- |
| Sex | F | M |  |
| **Clinical features** |  |  |  |
| Aphtous ulcers (vaginal/oral) | x | x |  |
| Thrush (oral/vaginal) | x | x |  |
| Recurrent respiratory infections | x | x |  |
| Hypothyreosis | x | x |  |
| Vitiligo | x |  |  |
| Lichen |  | x |  |
| Lymphadenopathy | x | x |  |
| Lymphoma |  | x |  |
| **Lymphocytes** (% of leukocytes) | 23 | 18 | **(% of parent)** |
| T cells | 74 | 51 | 66-88 |
| CD4+ | 55 | 48 |  |
| CCR7+CD45RA+ naive | 23 | 3 | 21-55 |
| CCR7+CD45RA- central memory | 50 | 35 | 8-33 |
| CCR7-CD45RA- effector memory | 24 | 55 | 20-52 |
| CCR7-CD45RA+ effector memory RA+ | 3 | 8 | 1-17 |
| CD38+HLADR+ activated | 7 | 9 | 1-5 |
| CD31+CD45RA+ recent thymic emigrant | 11 | 0,4 | 14-38 |
| CD8+ | 42 | 50 |  |
| CCR7+CD45RA+ naive | 16 | 2 | 19-71 |
| CCR7+CD45RA- central memory | 11 | 4 | 1-7 |
| CCR7-CD45RA- effector memory | 29 | 54 | 15-63 |
| CCR7-CD45RA+ effector memory RA+ | 44 | 40 | 4-34 |
| CD38+HLADR+ activated | 29 | 22 | 1-22 |
| CD31+CD45RA+ recent thymic emigrant | 12 | 0,5 | 17-65 |
| TCRgd | 1,4 | 0,7 | 2-12 |
| TCRab+CD4-cd8- | 1,4 | 0,2 | 3-9 |
| NK cells | 10 | 33 | 4-25 |
| B cells | 16 | 15 | 6-19 |
| CD27-IgD+ naive | 78 | 85 | 43-82 |
| CD27+IgD+ unswitched memory | 16,9 | 4,5 | 7,2-31 |
| CD27+IgD- class-switched memory | 1,9 | 8,4 | 6,5-29 |
| CD38lowCD21low CD21low | 18,8 | 2,9 | 1,1-6,9 |
| CD38++IgM++ transtitional | 7,7 | 11 | 0,6-3,5 |
| CD38++IgM- plasmablast/-cell | 0 | 1,2 | 0,4-3,6 |
| **Antibody levels** |  |  | **(g/l)** |
| IgG | 4,32 | 10,16 | 6,77-15,0 |
| IgG1 | 3,6 | 7,48 | 5,2-12,7 |
| IgG2 | 0,64 | 1,84 | 1,43-5,6 |
| IgG3 | 0,17 | 0,19 | 0,11-0,85 |
| IgG4 | <0,02 | 0,0003 | 0,03-2 |
| IgM | 0,22 | 1,82 | 0,88-4,84 |
| IgA | 0,59 | 0,69 | 0,36-2,59 |
| IgE | 3 | <1 | <1 |
| Vaccine responses | low | normal |  |

We performed whole exome sequencing (WES) for patients 1 and 2 but found no known PID-causing variants. Analysis of WES data filtered for rare variants shared by both patients (Suppl Table 1) identified a previously unreported heterozygous variant in *IKZF2* (chr2:213886829 G>T, NM_016260: c.C600A, p.Y200X) that introduces a premature stop-codon in the sequence coding for the fourth DNA-binding zinc-finger of HELIOS (Figure 1A). Targeted capillary sequencing validated the variant in the two patients and found all other tested relatives homozygous for the reference allele. The combined annotation-dependent depletion (CADD) score for this variant was 38, which is well above the mutation significance cutoff of 3.313 for *IKZF2* (Kircher et al 2014; Itan et al 2016). The position of this novel variant is highly evolutionary conserved (conservation score of 5.8, calculated using GERP++ (*20*)) further supporting that the variant is damaging.

**Fig 1.**
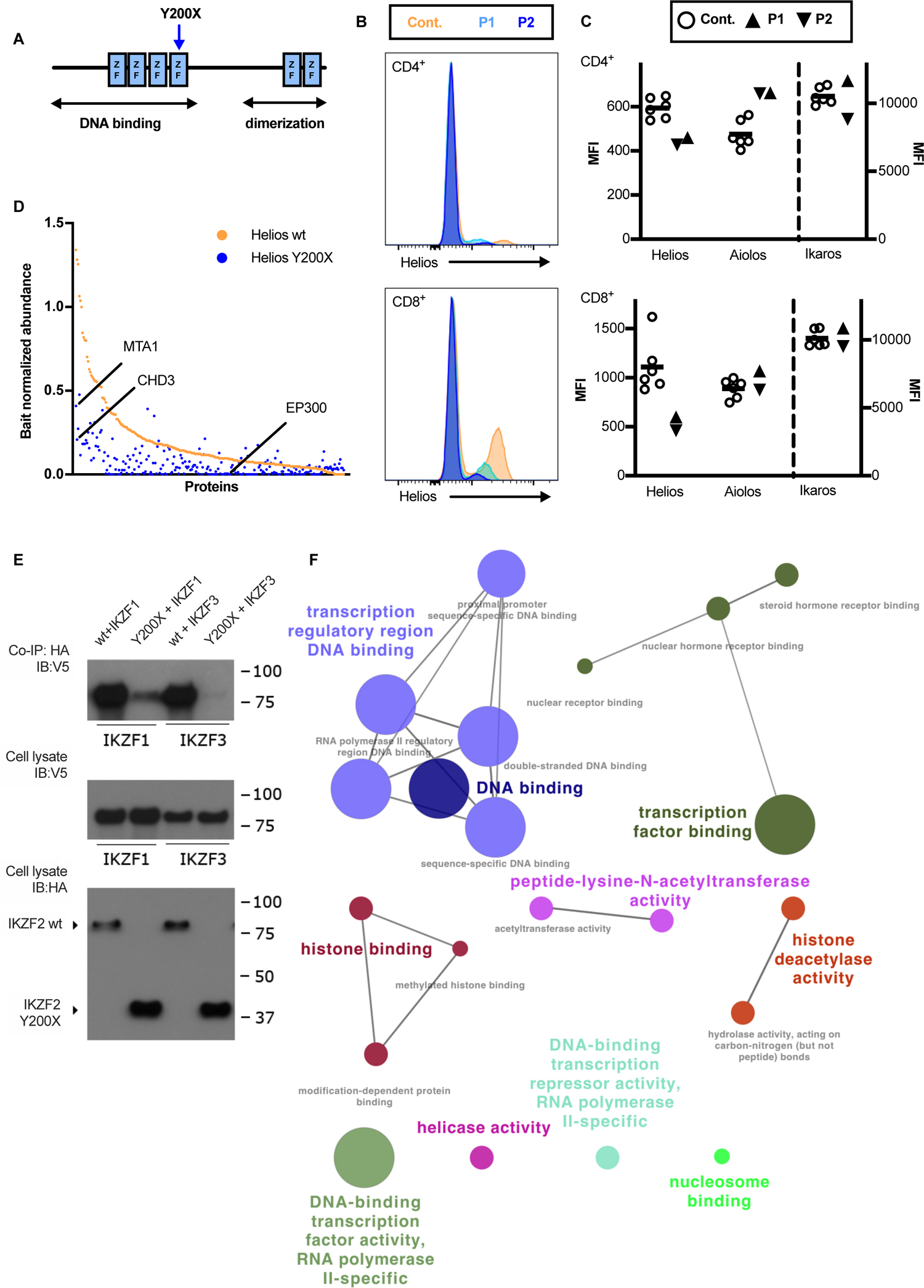
Truncating variant of *IKZF2* disrupts the protein-protein interactions of HELIOS. (A) Schematic representation of HELIOS protein. Blue arrow indicates the Y200X variant. (B) HELIOS expression in CD4^+^ and CD8^+^ T cells in patients compared to healthy controls. Histograms with patients and representative healthy control and (C) fluorescence intensity (MFI) of transcription factors HELIOS, AIOLOS, and IKAROS for patients and healthy controls in CD4^+^ and CD8^+^ T cells are shown. (D) Abundance of protein partners of wild-type (wt) HELIOS’ (orange) and variant Y200X (blue) HELIOS in interactome analysis. (E) A co-immunoprecipitation (Co-IP) assay displaying ability of HA tagged wild type HELIOS or variant Y200X HELIOS to form dimers with either IKAROS (IKZF1) or AIOLOS (IKZF3) tagged with V5. (F) The proteins whose protein-protein interactions were altered in Y200X compared to wild type HELIOS were selected and ClueGo clustering was performed to show the biological processes they were involved at. Cont.= control, P1=patient 1, P2=patient 2

Targeted capillary sequencing of the patient RNA derived cDNA indicated that the transcript containing the premature stop codon is not eliminated by the nonsense mediated RNA decay and may produce a truncated protein product (Supple Fig 1A). Based on these observations and previously reported functions of HELIOS in lymphocytes, we considered the *IKZF2* c.C600A, p.Y200X (here on p.Y200X for short) variant the most plausible candidate for further studies.

We evaluated the expression of HELIOS in patients and healthy controls by flow cytometry. HELIOS mean fluorescent intensity was considerably lower in patients in both CD4^+^ and CD8^+^ T cells compared with both healthy controls and unaffected relatives (Figure 1B, Suppl Fig 1C). Since IKAROS family members are known to form functional heterodimers between each other (*3*), we also quantified the expression of AIOLOS and IKAROS in patients. Mean expression of AIOLOS was higher in patients in total CD4^+^ helper T cells when compared with healthy controls. However, this difference was explained by a larger fraction of mature AIOLOS^high^ CD4^+^ T cells in patients (Suppl Fig 1D). No difference in IKAROS expression were detected (Figure 1C, Suppl Fig 1E). In summary, the novel *IKZF2* variant resulted in diminished HELIOS expression in affected individuals with no differences in expression levels of other IKAROS family members.

### Truncating variant abolishes HELIOS interactions with Mi-2/NuRD complex components

Since the variants premature stop-codon prevented the translation from the second zinc-finger domain onwards, we reasoned that it most likely affects the variant’s protein-protein interactions. In order to determine possible changes in interactions we performed biotin proximity ligation (BioID, (*21*)) in generated stable cell lines, expressing HELIOS constructs with MAC tag (Liu et al.) The variant p.Y200X lost protein-protein interaction with 48 proteins compared with wild-type HELIOS. Furthermore, interaction with 187 protein partners was significantly reduced compared to the wild-type HELIOS (Fig 1D and Suppl Table 2). Considering the central role of IKAROS family members as part of the Mi-2/NuRD complex (*22*), we specifically looked for altered interactions in this protein complex. The truncated HELIOS variant had reduced or lost interaction with 12 proteins involved in the Mi-2/NuRD complex, including Mi-2alpha and CHD3 that both are essential in the formation of the Mi-2/NuRD complex (*23*).

Since IKAROS family members heterodimerize strongly with each other, we tested if the variant p.Y200X affected HELIOS’ dimerization with IKAROS and AIOLOS. In a co-immunoprecipitation assay the variant’s dimerization with both AIOLOS and IKAROS was markedly impaired compared to the wild-type HELIOS (Fig 1E).

We used the ClueGo clustering tool (*24*) to identify the biological processes that were affected by variant’s lost protein-protein interactions. The proteins with altered interaction with the truncated HELIOS had functions predominantly linked to DNA modulation and transcription (ClueGO_MF, Suppl Table 2), and transcriptional repressors (ClueGO_BP, Fig 1F, Suppl Table 2). The interactome analysis thus indicated that the patient-derived *IKZF2* truncating variant impaired key HELIOS protein-protein interactions, including heterodimerization with AIOLOS and IKAROS.

### Increased T cell differentiation and augmented proinflammatory proteins in patients with the *IKZF2* p.Y200X variant

Immunophenotyping of T lymphocytes in the patients showed a decreased proportion of naïve CD8^+^ T cells with concomitant expansion of memory subsets. Especially the proportion of CD45RA^+^CCR7^-^ T effector memory RA^+^ (TEMRA) cells was higher in patients (Table 1). In CD4^+^ cells similar, though less pronounced, bias towards memory phenotype was observed. Patients also had increased number of activated CD38^+^HLADR^+^ cells *ex vivo* in both CD4^+^ and CD8^+^ T cell subsets (Table 1). In a more detailed flow cytometry immunophenotyping we could detect a skewing from naive T cells to effector memory - like phenotype that is also associated with T cell senescence (Fig 2A). Compared with healthy controls and non-affected relatives, a shift from naïve T cells to terminally differentiated effector memory cells was detected (Fig 2B). In all, the patients’ immunophenotype indicated chronic activation and increased maturation of T cells.

**Fig 2.**
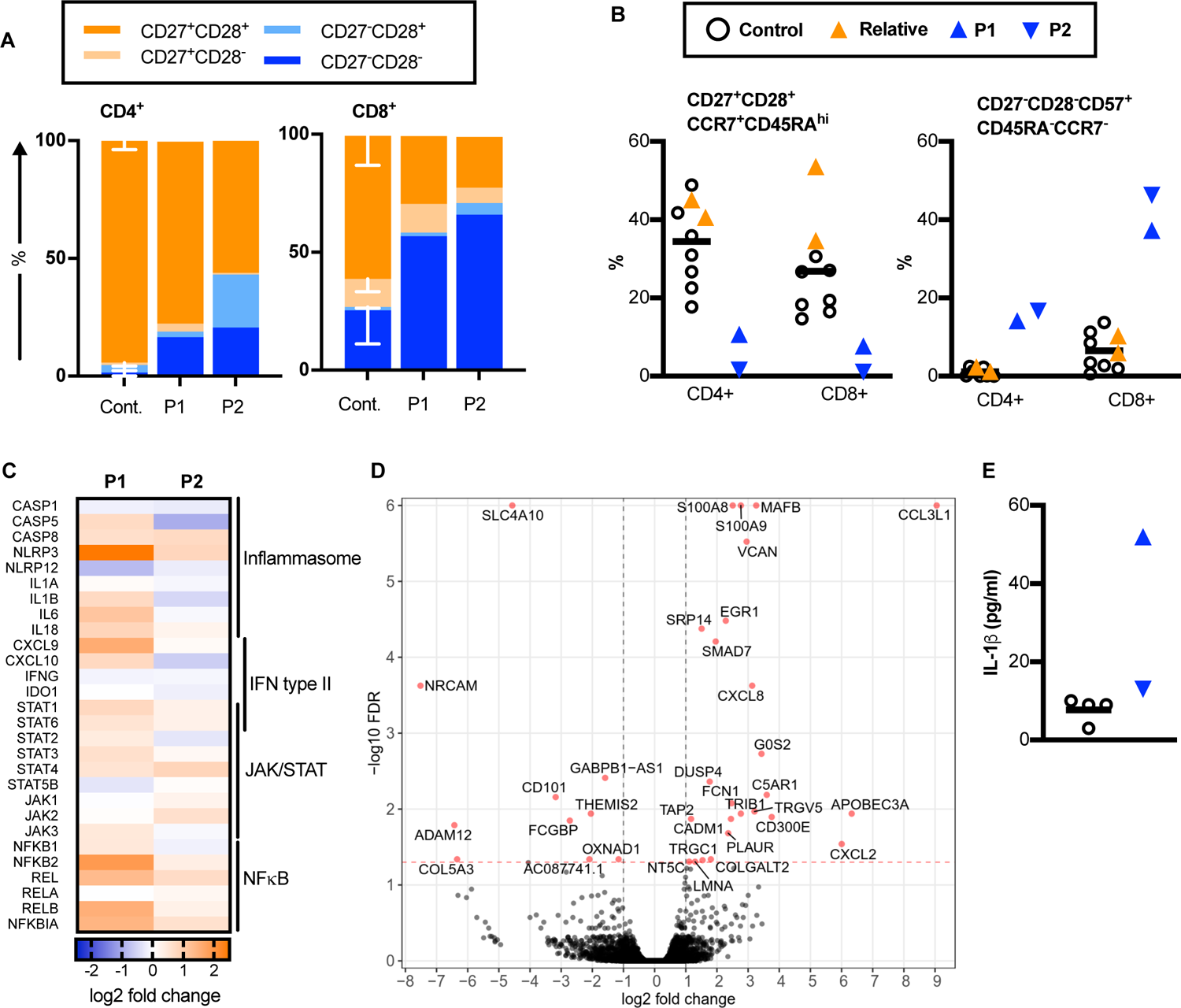
Chronically activated T cells in HELIOS haploinsufficient patients. (A) CD28 and CD27 expressing populations in patients and healthy controls is (n=6) is shown in CD4^+^ and CD8^+^ T cells, respectively. (B) Relative abundance of naive CD27^+^CD28^+^CD45RA^+^CCR7^+^ cells and terminally differentiated CD27^-^CD28^-^CD57^+^CD45RA^-^CCR7^-^ effector memory cells in patients and healthy controls and healthy relatives in CD4^+^ and CD8^+^ T cells, respectively. (C) Nanostring expression analysis of patients’ PBMCs showing differentially expressed genes compared to healthy controls. Log2 fold change to age and sex matched healthy control is shown. (D) Volcano plot of results from EdgeR analysis of CD8^+^ T cell 3’RNA-seq data. Orange dots represents genes showing significant (FDR<0.05) differential expression between patients and age and sex matched controls (n=3). (E) Levels of il-1β from culture supernatants containing unstimulated PBMC from patients and healthy controls, respectively. Cont.= control, P1=patient 1, P2=patient 2

In order to decipher the origin of the chronic activation in patients’ T cells, we used a custom gene panel (50 genes linked to immune signaling and inflammasome activation) with Nanostring nCounter (*25*) to screen for gene expression profiles in the patients’ peripheral blood mononuclear (PBMC) cells *ex vivo*. Both patients had a marked upregulation of genes related to both type 1 and type 2 interferon, NFkB, and JAK-STAT signaling (Fig 2C). Also a number of genes associated with inflammasome signaling were upregulated.

Cytotoxic CD8^+^ T cells had the highest HELIOS expression of effector T cell populations analyzed (Suppl Fig 2C&D) so we decided to do RNA sequencing (RNASeq) on magnetic bead enriched CD8^+^ T cells from patients and age and sex matched healthy controls to confirm and expand these findings from the Nanostring assay. In differential expression analysis using EdgeR several immunoregulatory and functional genes had altered expression in patients, such as upregulation of SMAD7 and downregulation of CD101 (Suppl Table 3). Pathway analysis using IPA showed that IFNgamma and IL1b downstream signaling pathways were markedly upregulated in patients (z-score 2.304, p=1×10-7 and Z-score 2.543 and P=6×10^-7 respectively, Suppl Table 4).

RNASeq showed that expression of genes S100A8 and S100A9 were upregulated in patients’ CD8 positive T cells *ex vivo* when compared with age and sex matched healthy controls (Fig 2D, Suppl Table 3). These genes code for a heterodimer called calprotectin that is a strong proinflammatory alarmin molecule. Calprotectin can be used as a biomarker for disease activity in many inflammatory conditions (*26*). To confirm the RNASeq result indicating increased calprotectin production, we measured the total calprotectin level from patientś sera. Patient 1 had 6.58 mg/l and patient 2 11.49 mg/l of total calprotectin in their serum, i.e. 1.2 and 2.1 times higher than the upper limit of reference values generated from healthy donors (*27*). The non-affected relatives had normal levels of calprotectin (1.95 and 3.72 mg/l).

Gene expression analysis of patients’ PBMCs revealed upregulation of genes relating to inflammasome activation. Calprotectin can activate inflammasome through TLR4 and induce il-1βeta production from target cells ((*28*)). We measured the serum levels of il-1βeta and found elevated concentrations from both patients: 1,95 pg/ml for patient 1 and 0,59 pg/ml for patient 2. All healthy controls were below the detection level of 0,31 pg/ml (n=21). Analysis of culture media of unstimulated PBMCs also revealed increased spontaneous secretion of il-1βeta for patients, although less pronounced for patient 2 (Fig 2E). However, no clear differences in inflammasome activation or cellular death were detected in comparison with healthy controls when patients’ monocytes were activated with LPS and combination of LPS and ATP (Suppl Fig 3A-C).

**Fig 3.**
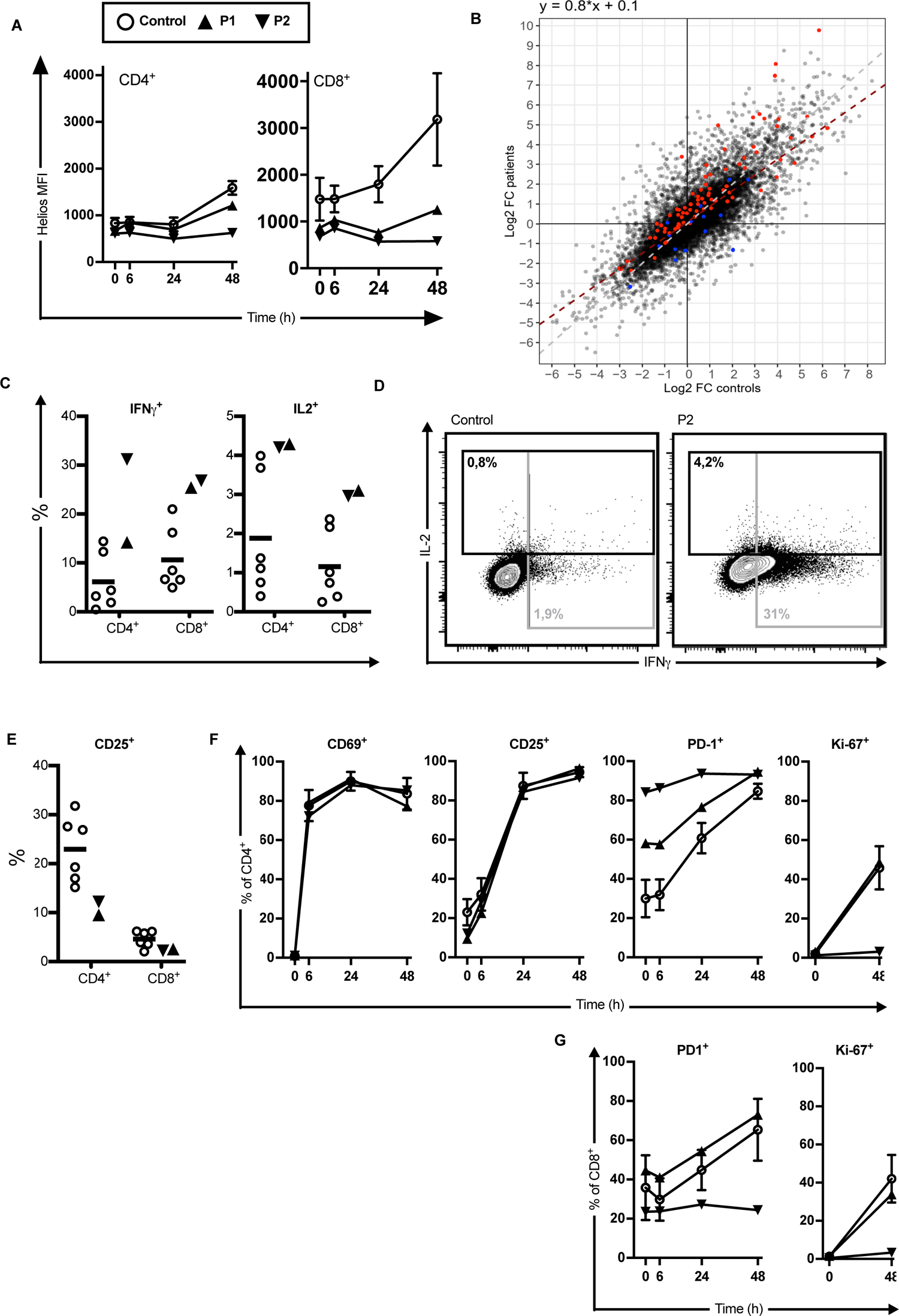
TCR activation leads to increased proinflammatory response in HELIOS haploinsufficient T cells. (A) Expression of HELIOS CD4^+^ and CD8^+^ cells, respectively, after TCR stimulation with immobilized anti-CD3-CD28 antibodies in patients and healthy controls. (B) Correlation in CD3^+^ T cell transcriptional regulation in response to 24 hour TCR stimulation between patients and healthy controls (n=5). Differential expression in unstimulated versus anti-CD3-CD28 stimulated cells and patients versus controls were analyzed from 3’RNA-seq data using EdgeR. Fitted linear regression is shown as a red dashed line (correlation coefficient 0.8) Genes found differentially upregulated or downregulated in patients versus controls in stimulated cells are marked in red and blue, respectively. (C) Percentage of both CD4^+^ and CD8^+^ T cells positive for IFNγ and IL-2, respectively, in patients compared to healthy controls after TCR stimulation and (D) dot plot of IFNγ and IL-2 expression in CD4^+^ cells of healthy control and patient 2 are shown. (E) Expression of IL-2 receptor alpha chain CD25 in CD4^+^ and CD8^+^ T cells ex vivo. (F) Relative abundance of cells expressing CD69, CD25, PD1 or Ki-67 in CD4^+^ T cells and (G) of cells expressing PD-1 or Ki-67 in CD8^+^ T cells in patients and healthy controls after TCR stimulation. P1=patient 1, P2=patient 2, (black triangles), MFI=mean fluorescence intensity. Controls = open circles.

These data indicate that the patients with the heterozygous p.Y200X variant, display a proinflammatory transcriptional signature both in their PBMCs and cytotoxic T cells. The increased proinflammatory milieu was also reflected by elevated serum levels of potent proinflammatory proteins.

### Increased IFNγ and IL-2 signaling in T cells from patients with the *IKZF2* p.Y200X variant

Both the maturation status of lymphocytes and ex vivo gene expression pointed to increased inflammatory signaling, so we proceeded to measure if the T cell receptor mediated activation was affected in patients. HELIOS is upregulated in response to TCR activation and currently there are no other known signaling pathways that control HELIOS expression (*15*). When we did a CD3-CD28 co-stimulation with immobilized activating antibodies on PBMCs *in vitro*, healthy controls showed a marked upregulation of HELIOS, especially in CD8^+^ T cells. However, with the same *in vitro* stimulation, our patients failed to upregulate HELIOS (Fig 3A).

Since TCR stimulation exacerbated HELIOS deficiency, we performed RNASeq on purified T cells from patients or age-sex matched healthy controls after 24 hr stimulation with immobilized anti-CD3-CD28 antibodies. We analyzed differential gene expression between patients and healthy controls in both unactivated and anti-CD3/CD28 activated CD3^+^ cells. Whereas only 10 genes were differentially expressed in the unactivated state (Suppl Fig 4E & Suppl Table 3), 103 genes showed differential expression after TCR activation including 16 downregulated and 87 upregulated genes (Suppl Fig 4F & Suppl Table 3). Analysis of sample pairs from the same individuals prior and after anti-CD3/CD28 activation also allowed us to compare transcriptional changes between groups in response to activation. As expected, a large percentage of expressed genes in CD3 cells were up- or downregulated in response to TCR activation (Suppl Fig 4G). To visualize the correlation in transcriptional regulation between patients and healthy controls, we plotted the gene-wise log2 fold changes of each group against each other (Fig 3B). Pearson’s correlation coefficient between patients and controls was 0.8, showing that by-and-large both groups exhibit similar up- and downregulation of gene expression in response to TCR activation. The genes differentially upregulated or downregulated in patients versus controls in the anti-CD3/CD28 treated cells are marked in red and blue, respectively. This illustrates that most of the differentially upregulated genes either retain higher expression or are upregulated more after TCR activation in patients than in controls.

**Fig 4.**
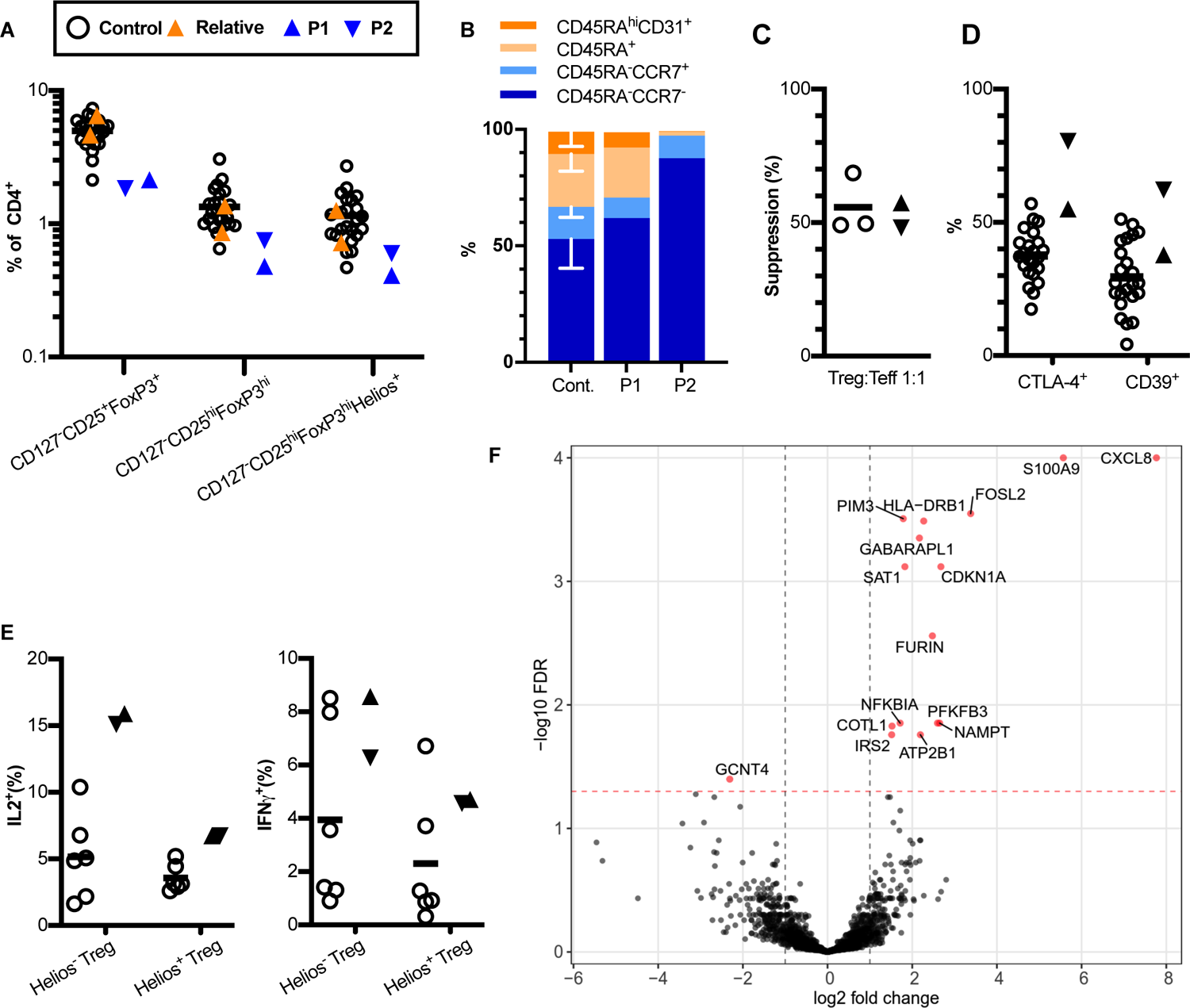
HELIOS haploinsufficient Tregs are activated and produce proinflammatory cytokines. (A) Proportion of regulatory T cells (Treg) of CD4^+^ cells gated as CD127^-^CD25^+^FOXP3^+^ to include also naive cells, CD127^-^CD25^hi^FOXP3^hi^ or CD127^-^CD25^hi^FOXP3^hi^HELIOS^+^ in patients, healthy relatives and healthy controls. (B) Relative abundance of recent thymic emigrant CD45RA^hi^CD31^+^, naive CD45RA^+^, and memory CD45^-^CCR7^+^ and CD45RA^-^CCR7^-^ cells in CD127^-^CD25^+^FOXP3^+^ Tregs in patients and healthy controls. (C) The suppression assay showing suppressive capacity of Tregs of patients and of healthy controls. (D) Expression of CTLA-4 and CD39 in CD127^-^CD25^+^FOXP3^+^ Tregs in patients and healthy controls. (E) Portion of IL-2^+^ and IFNγ^+^ cells, respectively, in both HELIOS^-^ and HELIOS^+^ Treg subsets in patients and healthy controls after overnight stimulation with immobilized anti-CD3-CD28 antibodies. (F) Volcano plot showing differentially expressed genes (orange dots) between patients and healthy controls from EdgeR analysis of 3’RNA-seq data from purified CD4^+^CD25^hi^CD127^-^ Tregs.

In pathway analysis of differentially expressed genes after CD3-CD28 activation both IFNγ and IL-2 downstream signaling was upregulated in patients (z-score 2,129, p-value 8×10^-10 and z-score 3,875, p-value 8×10^-13, respectively; Suppl Table 4). Also the T cell exhaustion transcriptional signature (z-score 0,813, p-value 1×10^-4) and NFkB signaling pathway (z-score 1,342, p-value 0,005) were upregulated compared with healthy controls. When we cross-analyzed the protein partners whose interaction with the variant HELIOS was affected, we could detect changes in downstream signaling of several of HELIOS’ protein partners (Suppl Table 4). Taken together, the transcriptional landscape after TCR stimulation in patients with the p.Y200X variant of IKZF2 compared with healthy controls indicated immune dysregulation of several major immune activation pathways.

Both Nanostring analysis on PBMCs and RNASeq T cells *ex vivo* and after stimulation suggested increased IFNγ signaling. Another important pro-inflammatory cytokine controlled by Helios is IL-2 (*12*) so we analyzed the IFNγ and IL-2 production in T cells after TCR stimulation. Increased proportion of IFNγ - and IL-2 - producing cells was observed in patients in response to TCR stimulation (Fig 3C&D). IL-2 receptor alpha chain (CD25) expression levels were lower *ex vivo* in patients compared to controls (Fig 3E). Low CD25 could result from downregulation of the IL-2 receptor in response to higher baseline level of IL-2. We also measured the soluble CD25 from patients’ sera and it was within reference values (Suppl Clinical data). The kinetics of activation marker expression after TCR stimulation on both CD4 and CD8 effector T cells were comparable between patients and healthy controls indicating that reduced HELIOS expression does not affect the overall expression of activation markers on T cells (Fig 3F&G). One patient had significantly reduced proliferative response of both CD4^+^ and CD8^+^ T cells to TCR stimulation as measured by both CFSE assay and failed upregulation of Ki67 (Fig 3F&G).

To summarize, reduced expression of HELIOS in T cells is associated with altered response to TCR stimulation. More specifically, we could detect increased production and downstream signaling of proinflammatory cytokines in effector T cells.

### Patients with the *IKZF2* p.Y200X variant have an increased proportion of T regulatory cells with a proinflammatory phenotype but normal suppressive activity

Helios expression is high in Tregs and it stabilizes Treg suppressive function in mice (*10*) so we next measured if the Tregs from patients also produced more proinflammatory cytokines similar to what has been reported in Helios-deficient mice (*12*). In our patients, the proportion of circulating naive and activated Tregs was lower than in healthy controls or unaffected relatives (Fig 4A). More detailed characterization of Tregs revealed a shift from naive Tregs to more mature phenotype in patients as in T cells in general. Proportion of recent thymic emigrant Tregs was also lower in patients (Fig 4B). However, the suppressive function of Tregs *in vitro* was not affected in suppression assay (Fig 4C). This apparent discrepancy could be partly explained by the high expression level of immunosuppressive protein receptor CTLA-4 and suppressive adenosine producing ectonucleotidase CD39 (Fig 4D). Patients’ Tregs were thus more mature and activated as compared with healthy controls which could in part compensate for the reduced numbers.

*In vitro* Treg suppression assays have several limitations and do not reflect all functional aspects of Tregs so we continued to do a transcriptome analysis of Tregs. We sorted CD127^-^ CD25^hi^ T cells from patients and age and sex matched healthy controls (Suppl Fig 5C). The changes in transcriptome were less pronounced than in effector T cells. Treg analysis was somewhat limited by the low number of Treg cells and RNA yield. As in effector T cells the IFNγ and NFkB signaling were higher in patients (z-score 2,271, p-value 9×10^-9, and z-score 1,623, p-value 2×10^-4, respectively). These most likely reflect the overall proinflammatory milieu in the patients. However, some Treg-specific changes could be seen. For example the paraprotein convertase Furin expression was higher in patient Tregs (Fig 4F). Furin has been reported to be important for the suppressive capacity of Tregs and it is upregulated in the activated Tregs (*29, 30*). Together, Furin, CD39, and CTLA-4 upregulation indicate that patients’ Tregs were more activated than in healthy controls (Suppl Fig 5B).

**Fig 5.**
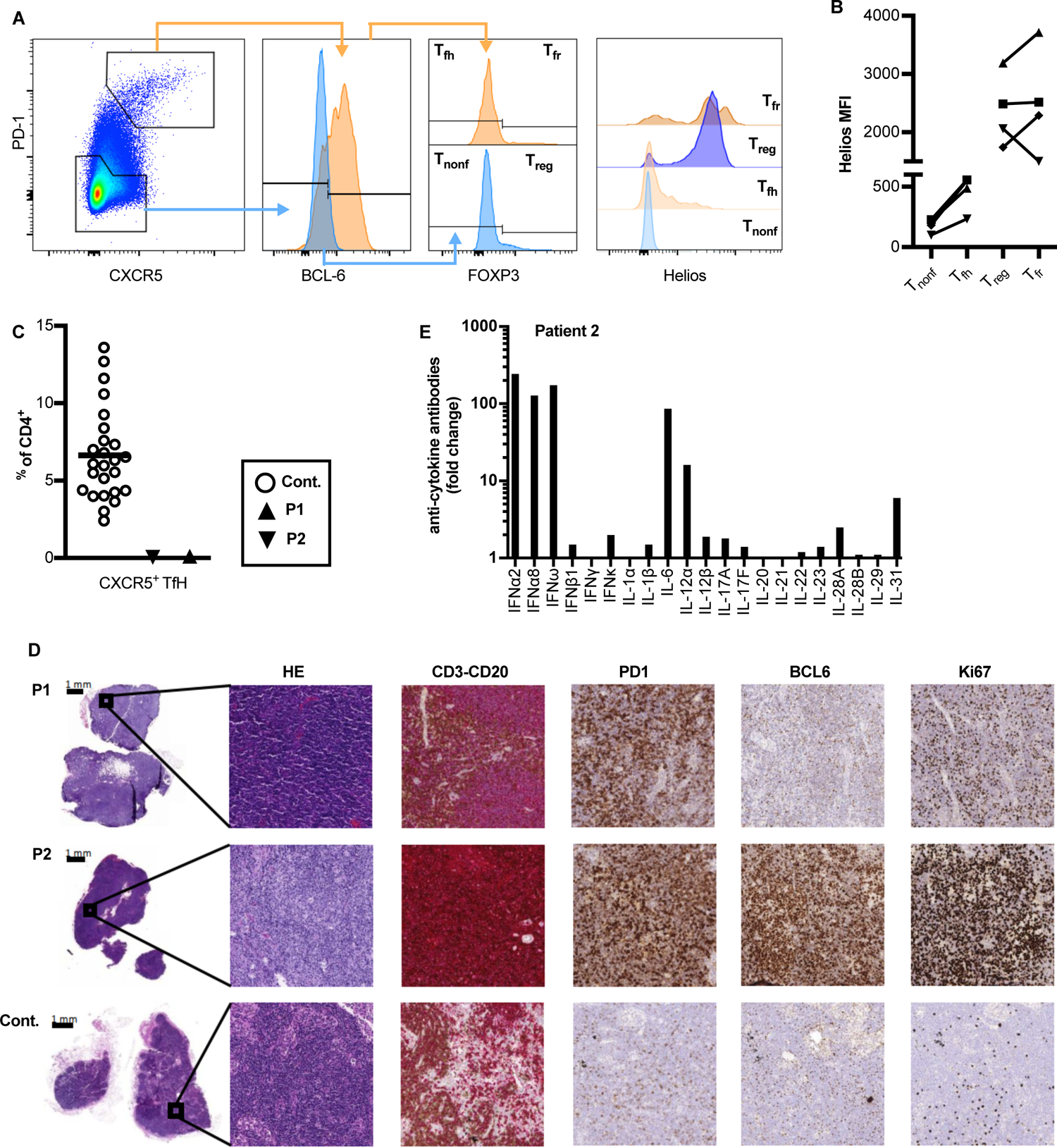
Accumulation of Tfh cells in the perifollicular areas of lymph nodes. (A) HELIOS expression in lymph node CD3^+^CD4^+^ T cell was analyzed in FOXP3^-^CXCR5^low^PD1^low^BCL-6^-^ T non-follicular helper (Tnonf), CD4^+^FOXP3^-^CXCR5^hi^PD1^hi^BCL-6^+^ T follicular helper (Tfh), CD4^+^FOXP3^+^CXCR5^low^PD1^low^BCL-6^-^ T non-follicular regulatory (Treg) and CD4^+^FOXP3^+^CXCR5^hi^PD1^hi^BCL-6^+^ T follicular regulator (Tfr) cells. Gating strategy in a healthy control and (B) mean fluorescence intensity (MFI) of HELIOS for four individual donors is shown. (C) Frequency of CD4^+^ T cells expressing follicular homing receptor CXCR5^+^ in PBMCs of patients and healthy controls. (D) Histological analysis of patients’ lymph nodes showing follicular hyperplasia in the perifollicular region around the germinal centers. A more detailed immunohistochemistry staining of accumulated cells expressed with Tfh markers Bcl6 and PD-1 and Ki-67 indicating proliferation activity. (E) Auto-antibodies against cytokines in patient 2. Fold change compared to mean of healthy controls (n=5) is shown. Cont.= control, P1=patient 1, P2=patient 2

Helios-deficiency in mice has been reported to lead to proinflammatory cytokine production in Tregs mice (*10*) so we proceeded to test how fresh Tregs isolated from our patients responded to stimulus. After *in vitro* TCR stimulation the patients’ Tregs had a higher proportion of IL-2 producing Treg cells but there was no clear difference for IFNg production (Fig 4E). Since diminished IL-2-STAT5 signaling has been linked to Treg instability in Helios^-/-^ mice (*10*) we also evaluated STAT5 phosphorylation in response to IL-2 stimulation. No differences were observed (Suppl Fig 5D). These data indicate that Tregs in patients with heterozygous loss-of-function variant in *IKZF2* are skewed towards a more activated and mature phenotype. These Tregs retain their suppressive capacity but also produce more IL-2.

### Dysregulated germinal center reactions and aberrant antibody production in patients with the ***IKZF2* p. Y200X variant**

Both patients’ clinical presentation with recurrent upper and lower respiratory infections suggested common variable immunodeficiency - like B cell pathology. Clinical immunophenotype of the B cells revealed an increased proportion of transitional B cells in both patients. Patient 1 also had a higher number of activated B cells, but decreased amount of switched memory B cells and plasmablasts in line with hypogammaglobulinemia. Since HELIOS expression in B cells is low (Suppl Fig 2B and 3C) we reasoned the B cell defect could result from impaired T cell help to B cells. Supporting this assumption, Helios knockout mice have defective regulatory follicular T cells (Tfr) with accumulation of Tfh cells to the lymph node and aberrant germinal center formation (*11*). Moreover, in humans HELIOS has been suggested to have a role in the Tfh cell differentiation *in vitro* (*17*). To evaluate the role of HELIOS in human Tfh and Tfr cells we measured HELIOS expression in these subsets isolated from fresh lymph node samples obtained from organ donors. HELIOS expression in Tfr cells was comparable to non-follicular Tregs (Fig 5A&B). In Tfh cells HELIOS expression was lower than in Tfr but higher when compared to non-follicular effector T cells (Fig 5A&B). These data support a role for HELIOS in the follicular effector and regulatory T cell populations.

In peripheral blood, T cells positive for the homing marker CXCR5 and high PD1 expression form a population enriched for circulating Tfh cells. Their number was markedly lower in patients’ peripheral blood (Fig 5C) and we could not reliably detect circulating CD4^+^FOXP3^+^CXCR5^+^ regulatory follicular T cells from patients while in controls they accounted for 0,23 (+/- 0,11) % of CD4^+^ cells.

Since Tfh cells carry out their effector functions in lymph nodes and preanalytical factors may affect reliable CXCR5 detection, we performed immunohistochemical analyses on archival resected lymph nodes from both patients. The lymph nodes were removed due to persistent lymphadenopathy and were non-malignant. Pathologic-anatomic diagnosis on both was follicular hyperplasia, a common unspecific finding in a variety of diseases, including autoimmunity. In a more detailed immunohistochemical analysis both patients had increased CD3^+^ cellularity in the perifollicular region. These cells expressed high levels of Bcl6 and PD1 so they most likely represent an accumulation of Tfh cells in the perifollicular region around the germinal centers (Fig 5D). We also detected an increased proliferative activity as indicated by the high Ki67 labeling index.

The lack of circulating Tfh cells and accumulation of Tfh-like cells in the light zones of the lymph nodes suggest a dysregulated germinal center reaction that often leads to production of autoantibodies. Both patients were at the time of clinical examination negative for anti-nuclear autoantibodies and anti-thyroid autoantibodies even though both had hypothyroidism (Suppl Clinical Data). We next measured neutralizing antibodies against cytokines because these autoantibodies have been reported in a number of immune dysregulatory conditions (*31*). Patient 2 had high titers of antibodies against multiple cytokines, especially type 1 interferons (Fig 5E). This anti-cytokine autoantibody profile was reminiscent of what is commonly seen in APECED patients (*32*) which is a syndrome of severe immune dysregulation. Patient 1 had hypogammaglobulinemia and also her pneumococcal vaccine responses were impaired (Table 1 & Suppl Clinical Data) so autoantibody measurements were unreliable. Anti-cytokine antibody titers against few cytokines were slightly higher in patient 1 sera compared to controls but not in the magnitude exhibited by patient 2 (Suppl Fig 6B).

**Fig 6.**
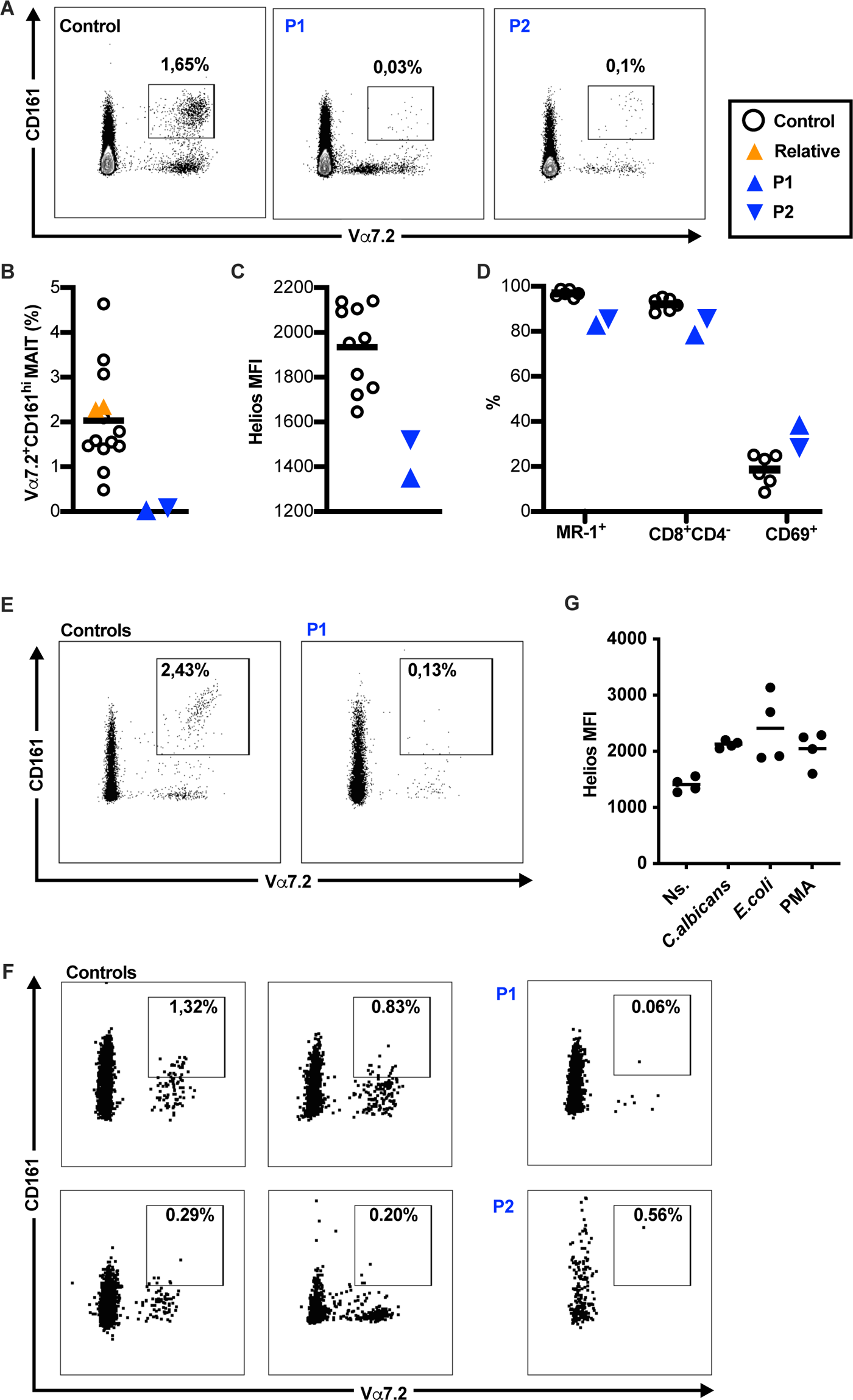
Reduction of circulating MAIT cells. (A) Dot plots showing abundance of Vα7.2^+^CD161^+^ MAITs in the CD3^+^ T cells from peripheral blood in patients and representative healthy control and (B) their frequency in CD3^+^ T cells in patients, healthy relatives and healthy controls. (C) Mean fluorescence intensity (MFI) of HELIOS in MAITs. (D) Abundance of MR1^+^, CD4^-^CD8^+^ and CD69^+^ cells, respectively, in MAITs in patients and healthy controls. (E) Frequency MAITs in colon and (F) in duodenum samples obtained from patients and organ donors without any chronic inflammatory conditions. (G) HELIOS expression in MAITs from PBMC in response to stimulation with *E. coli, Candida albicans* or PMA.

We can conclude that patients with heterozygous p.Y200X variant in *IKZF2* had abnormal antibody findings - hypogammaglobulinemia and anti-cytokine autoantibodies - together with signs of T follicular helper cell dysregulation in the lymph nodes.

### MAIT cells are reduced in circulation and in gut epithelium of patients with the *IKZF2* **p.Y200X variant**

Innate lymphoid cells including Natural Killer cells (NK) have been reported to express HELIOS (*33*) but NK cell immunophenotyping from the patients did not show any significant perturbations in NK cell subpopulations (Suppl Fig 7). MAIT cells are innate like T cell subset contributing to bacterial and fungal defense on mucosal surfaces and have a high expression level of HELIOS (*34*). Since both patients had mucosal *Candida albicans* infections and recurrent bacterial infections, we next decided to analyze their MAITs. In comparison to healthy controls and unaffected relatives, the number of Valfa7.2^+^CD161^hi^ T cells in circulation was markedly reduced in patients (Fig 6A&B). The small number of Valfa7.2^+^CD161^hi^ T cells detected in patients also had lower HELIOS expression than healthy controls (Fig 6C). The TCR Valpha7.2 positive and CD161 high population is enriched for MAITs but it also contains conventional T cells. To more specifically measure MAIT number in our patients, we used the MR1 tetramer loaded with 5-(2-oxopropylideneamino)-6-D-ribitylaminouracil (5-OP-RU) that is specific for the invariant TCR found in majority of MAITs (*35*). Only 84% of patients’ Valfa7.2^+^CD161^hi^ cells were positive for 5-OP-RU loaded MR1 tetramer compared to 97% in the controls (Fig 6D). Also, the proportion of CD8^+^CD4^-^ MAITs, proposed to represent a more active phenotype (*36*), was lower in patients (Fig 6D). The expression of activation and tissue retention marker CD69 was higher in patients’ MAITs (Fig 6D). In all, the number of MAITs was reduced in patients’ circulation and the remaining small Valfa7.2^+^CD161^hi^ population contained more conventional T cells than that of healthy controls.

Reduced MAIT number in the blood can be caused by their recruitment to sites of inflammation in the periphery as seen in e.g. inflammatory bowel diseases (*37*). We measured MAITs by flow cytometry from intestinal biopsies taken from our patients during clinically indicated gastroscopies and a colonoscopy. Both patients had unspecific gastrointestinal complaints but no diagnostic findings were made in the endoscopies. We analyzed duodenal biopsies from both patients and a colon biopsy from patient 1. We found a low frequency of Valpha7.2^+^CD161^hi^ cells in mucosal samples in both patients. The proportion of MAIT-like cells isolated from mucosal biopsies was not higher when compared with samples obtained from organ donors (Fig 6E-F). Therefore, the low proportion of MAITs in circulation appeared not to be a result of an increased accumulation to the gut mucosal sites.

The role of HELIOS expression in MAITs is unknown. MAITs develop in the thymus as a minor population. They expand and acquire effector memory - like phenotype quickly after egressing from the thymus (*38*). We wanted to evaluate at which stage of their development and function MAITs start to express high levels of HELIOS. We analysed the expression of HELIOS in developing MAITs isolated from thymic samples (n=4) acquired from children undergoing cardiac surgery. As reported before, the percentage of developing MAITs was low and only 0,025% (+/- 0,015%) of thymocytes were positive for the MR1-tetramer. The majority of MR1-tetramer positive thymocytes expressed HELIOS (93,5% +/- 5,6%, Suppl Fig 8D). Next, we used stimulation with *E. coli* and *C. albicans* on PBMCs (*39*) from healthy donors to measure if HELIOS is involved in the peripheral activation of MAITs. Unstimulated MAITs were already predominantly HELIOS positive but both microbial stimulations caused a robust upregulation of HELIOS in MAITs (Fig 6G). We can therefore conclude that HELIOS is expressed both during the thymic development of MAITs and their activation in the periphery.

## Discussion

Here we describe a novel immunodeficiency with signs of immune dysregulation caused by heterozygous germline loss of function mutation in *IKZF2* coding for the transcription factor HELIOS. The truncating mutation found in this single pedigree profoundly changed the ability of HELIOS to interact with proteins of the NuRD complex and other members of the IKAROS family. Affected patients had a reduced level of HELIOS expression in their peripheral T cells and specific immunophenotypic changes in T cell populations expressing high levels of HELIOS, especially MAIT cells. The immunophenotype segregated in the pedigree with the carriers of the loss-of-function *IKZF2* variant.

Protein interactome analysis of the truncated protein variant showed that it lost interaction with key components of the NuRD complex, the protein complex in which IKAROS family members are involved in chromatin remodelling (*3*). RNAseq of stimulated primary patient lymphocytes confirmed altered gene expression directly downstream of some of the transcription factors identified with the interactome analysis. For example, downstream signaling for histone acetyltransferase EP300, that we found to interact only with wild type but not with the variant protein, was altered in all three RNAseq conditions (Suppl Table 2&4). Based on our findings, the interactions with the NuRD complex are essential for HELIOS’ function.

Helios acts mainly as a transcriptional repressor in lymphocytes (*3, 4*). In accordance with this, we detected increased proinflammatory signaling and production of proinflammatory cytokines and proteins in the carriers of the loss-of-function variant. The chronic overactivation of the immune system is most likely the cause for the patients’ activated mature T cell - skewed immunophenotype. Our detailed analysis of patients’ immune system concurs with earlier data acquired in murine models, but also offers new insight into the role of HELIOS in regulating immune responses. Helios represses IL-2 production (*12*). In our patients, we saw enhanced IL-2 production from both effector and regulatory T cells. Also the transcriptome analysis of T cells indicated increased IL-2 signaling. Interestingly, patients’ cytotoxic T cells produced higher amounts of calprotectin, which is an alarmin molecule responsible for e.g. sterile inflammasome activation. Calprotectin is used in various clinical settings to evaluate subclinical systemic inflammation (*40*). Concomitantly the patients had increased levels of IL-1β cytokine, reflecting the inflammasome activation.

The studies on the role of Helios in controlling immune responses have mainly concentrated around its effects on regulatory T cells. Selective Helios knockdown with the FOXP3 promoter is sufficient to induce autoimmunity in mice (*10, 11*). On the other hand, in Helios^-/-^ mice suppressive capability of Tregs *in vitro* is intact and their *in vivo* function is only mildly impaired (*11, 41*). Our patients had only mild autoimmune manifestations and the Treg *in vitro* suppression assay did not indicate any reduction in suppressive capacity. Moreover, patients’ Tregs were skewed towards a more mature immunophenotype with high CTLA-4 expression and signs of activation, which could indicate some compensatory mechanisms making up for the reduced HELIOS expression. Helios has been suggested to stabilize the suppressive phenotype of Tregs (*12*) and a loss of Helios could cause Treg conversion to effector T cells (*13*). Patients’ Tregs had a more inflammatory phenotype with increased IL-2 production and lower FoxP3 and Helios expression, which is characteristic of unstable Tregs (*13, 42*),. Our results confirm earlier reports that HELIOS has a limited role in stabilizing human Tregs.

Germline knockdown of Helios or selective knockdown of Helios with FOXP3 promoter increases germinal center formation and accumulation of Tfh and germinal center B cells in lymph nodes (*11*). In patients’ lymph nodes we could see an accumulation of perifollicular T cells that are considered the precursor population for mature Tfh cells. Similar increase of Tfh cells in lymph nodes after immunization was evident in Helios heterozygous mice (*11*). Circulating PD1^hi^CXCR5^+^ Tfh cells are fully mature memory cells that have exited from the germinal centers (*43*). In our patients, the circulating CXCR5 positive Tfh cells were almost undetectable. It is thus possible that HELIOS is required for the terminal Tfh differentiation within the germinal center. Another plausible explanation is that HELIOS is somehow involved in the Tfh egress to circulation from the germinal center.

One other defining feature of our patients was the marked reduction in the number of MAITs in the peripheral blood. This could be a result of increased homing of MAITs to mucosal tissues but analysis of mucosal biopsies from the patients detect only a small fraction of MAITs. Both patients suffered from recurrent mucosal *Candida albicans* infections which could be partly explained by the loss of MAITs from the mucosa that are important effectors in fungal infections (*44*). Deficiency of HELIOS could also harm the MAIT development in the thymus or their peripheral activation and expansion. Since the patients had so few MAITs we could only perform experiments on MAIT biology in cells from healthy donors. Based on our findings in the healthy human thymus we can conclude that HELIOS is highly expressed already in developing MAITs. However, HELIOS expression was also high in mature effector MAITs and even further upregulated after microbial stimulation. Thus, although our data is inconclusive on at which stage the loss-of-function variant in *IKZF2* could affect the MAIT numbers, our data supports a non-redundant role for HELIOS in MAITs. An alternative explanation is that the chronic inflammation in patients caused the MAIT cell depletion. MAIT cell numbers have been shown to have an inverse correlation with levels of innate proinflammatory cytokines in various inflammatory conditions (*45, 46*).

Clear limitation of our study is the small number of patients from a single family. This is bound to affect e.g. RNASeq analyses since some of the differences we detected could be caused by other inheritable factors than the *IKZF2* variant. However, our results from these two patients recapitulate the main findings from Helios knock-out mice – IL-2 producing Tregs and dysregulated germinal centers. In addition to HELIOS’ known effects on the immune system, we show that HELIOS has a previously underappreciated role in the MAIT cell lineage. Further studies are needed to understand what the functional significance of HELIOS is both in the control of germinal center reactions and MAITs. Heterozygous missense mutations in IKAROS cause CVID-like immunodeficiency with progressive loss of B cells in the circulation with variable penetrance of the clinical disease (*7*). Dominant negative mutations in IKAROS lead to an early onset combined immunodeficiency phenotype with disturbed T cells effector maturation and dysfunctional monocytes (*8*). These variable immunological presentations highlight the complex roles IKAROS transcription factors have as activators and repressors of transcription in the immune system. Shahin et al. show that homozygous missense mutation in HELIOS leads to combined immunodeficiency with hypofunctional T cells while the truncating variant in our patients leads to overactivation of the immune system with relatively mild immunodeficiency. These different presentations emphasize how highly variable functions HELIOS has in T cells.

## Materials and methods

### Study subjects and samples

The study was conducted according to the principles of the Declaration of Helsinki. The study was approved by the ethics committee of Helsinki University Hospital (138/13/03/00/2013 and HUS/747/2019) and written informed consent was obtained from participants. The control group for blood samples consisted of 25 healthy individuals aged 24-66 (mean 46) years, 14 of them female. In addition samples were acquired from two relatives of the patients without mutation in *IKZF2* gene. The number of controls varied among experiments and it is indicated either in the figures or in the text. In experiments containing less than 10 healthy controls the controls were sex and age matched. Patients have approved the publication of the manuscript.

Patients’ clinical T- and B-cell phenotype, total blood count, immunoglobulin levels, complement activity and anti-tetanus and -diphtheria antibodies were evaluated by clinically validated test in Tampere University Hospital’s clinical laboratory Fimlab (Tampere, Finland).Samples from duodenum and colon were acquired from the patients during diagnostic endoscopy. Lymph node samples from patients were archival diagnostic samples. Control tissues samples were obtained from organ donors: duodenum from four individuals (34-69 years, 2/4 female) and colon from one individual (a female in her 40s), and 4 lymph node samples for characterization of follicular T cells (agerange 19-41, all male). The human thymi were obtained from four children undergoing cardiac surgery ( three infants, one teen, 3/4 females).

### DNA extraction and sequencing

Genomic DNA was extracted from EDTA blood with the Qiagen FlexiGene DNA kit (Qiagen) or from Oragene OG-575 saliva collection kit with the prepIT-L2P kit (DNA Genotek). For RT-PCR, RNA was extracted with the Qiagen miRNeasy kit (Qiagen) from freshly isolated PBMCs, and reverse transcribed into cDNA using SuperScript™ VILO™ cDNA Synthesis Kit (ThermoFisher). Exome and capillary sequencing were performed at the sequencing core facility of the Institute for Molecular Medicine Finland (FIMM). Exome libraries were generated using the Clinical Research Exome (Agilent Technologies) or the Nextera Flex (Illumina) capture kits, and sequencing was performed with 101 bp read length on the HiSeq1500 or the NovaSeq6000 Sequencing Systems (Illumina) or the NovaSeq 6000 Sequencing System (Illumina), respectively. Read mapping and variant calling were performed with an in-house pipeline. Variant annotation was performed with ANNOVAR(*47*). Variant data was filtered following a dominant inheritance model as described in Suppl Table 1. For frequency filtering, we utilized population level variant frequency data from the Genome Aggregation Database (gnomAD) (*48*) and 1000 genomes(*49*). Recurrent sequencing artefacts and bad quality variants were excluded based on in-house data and visual inspection of reads on the Integrative Genomics Viewer(*50*). Candidate variants were validated by targeted PCR by DreamTaq Green PCR Master Mix (ThermoFisher)and capillary sequencing on the ABI3730XL DNA Analyzer (Applied Biosystems). All primers used for PCR and capillary sequencing are listed in Supplemental Table 5.

### Protein-protein interaction analysis

Biotin proximity ligation assay was done with stable cell lines, generated from Flp-In™ T-REx™ 293, expressing HELIOS constructs with N-terminal MAC tag ((*51*). Cell line generation, sample preparation and mass spectrometry was done as previously described (*51*), with the exception of using 1% N-dodecyl maltoside instead of 0.5% IGEPAL in the lysis and wash buffers.

Peptides with a false discovery rate (FDR) of <0.05, were exported from the peptides detected with mass spectrometry. Identified proteins were compared against the Contaminant repository for affinity purification database (*52*). Only interactions with <20% frequency and 2-fold higher abundance, compared to the corresponding protein values in the controls database, where classified as high-confidence interactions. The Cytoscape software platform (*53*) was used to visualize the high-confidence protein-protein interactions. ClueGo plugin (*24*) was utilized for clustering of biological processes the identified proteins contributed to.

### Co-immunoprecipitation

6*10^5 HEK293 cells were plated in 6-well plate wells and transfected on the next day. Each was co-transfected with 1 μg of either HA tagged wild type IKZF2 or C600A IKZF2 and V5 tagged IKZF1 or IKZF3. 24h after transfection, cells were washed with cold PBS, lysed on ice for 15 min in 1 ml of ice-cold lysis buffer (0.5% IGEPAL, 50mM HEPS, 5mM EDTA, 150mM NaCl, 50mM NaF pH 8.0 supplemented with 1mM DTT, 1mM PMSF, 1,5mM NaVO_4_, 1x Sigma protease inhibitor cocktail). Lysates were pipetted into fresh 1.5 ml tubes and centrifuged 16 000g, +4°C for 15 min to remove insoluble debris. 20 μl of supernatants were taken into fresh tubes with 20 μl 2x laemmli sample buffer and incubated at 95°C for 5 min (lysate sample). 950 μl of supernatants were moved to new tubes with 30 μl of washed anti-HA beads (A2095, Sigma), and incubated 2h on rotation in +4°C. Beads were spinned down and the supernatant was discarded. The beads were washed three times with 1 ml of ice-cold lysis buffer. The immunoprecipitated proteins were eluted with a 2x laemmli sample buffer and incubated at 95°C for 5 min and the beads were spinned down (Co-IP sample).

### Western blot analysis

Co-immunoprecipitation (10 μl) and lysate (5 μl) samples were loaded on precast SDS-PAGE gels (any kD gel with 15 wells, Mini-Protean TGX, Bio-Rad) and transferred onto nitrocellulose membrane (NBA085C001EA, PerkinElmer) with semi-dry transfer (Trans-blot SD semi-dry transfer cell, Bio-Rad). The membranes were blocked with 5% milk – in 0.05% Tween – TBS. HA antigen was detected with primary HA antibody (16B12 Biolegend, 1:2000 dilution in blocking solution), V5 antigen was detected with primary V5 antibody (R960-25 Invitrogen, 1:5000 dilution in blocking solution). Primary antibodies were detected with a secondary antibody coupled to HRP (NA931 GE, 1:1000 dilution in blocking solution). ECL reaction (RPN2232, Amersham) was developed on photographic films (Super RX-N, Fuji X-ray films).

### Sample preparation

Blood was drawn into Li-heparin Vacutainer tubes (BD Biosciences), plasma was separated by centrifugation, and PBMCs isolated using Ficoll-Paque (GE Lifesciences) gradient centrifugation. The cells were cryopreserved using CTL-Cryo ABC (CTL) kit. Cryopreserved samples were used unless otherwise stated.

Sample resecates from duodenum, lymph nodes and thymus were wrapped in tissues dampened with cold saline solution and kept on ice. Duodenal biopsies were transported in full media and kept on ice. Immune cells from fresh tissues were extracted within a maximum of 6 hours of operation and analyzed subsequently. The duodenal samples were cut in small pieces, rinsed with PBS and incubated in 5 mL of enzyme solution (RPMI containing 15 mM HEPES, 0.25 mg/mL of DNAse I and 0.25 mg/mL of collagenase II) on a magnetic shaker in +37°C water bath for 20-30 min. The tissue digest was filtered with 100 micron filter and washed first with cold PBS containing 10% FCS or human AB media. Lymph nodes were identified from ileal mesenterium and excess adipose tissue was removed. Lymphocytes were mechanically extracted using a 40 micron filter and a plunger. Thymocytes were released with mechanical homogenisation of the thymic resecate. All single cell sample solutions were washed 2 times with Staining buffer (PBS containing 2% FCS and 2 mM EDTA) before use in downstream applications.

### Flow cytometry

For staining of surface antigens fresh or thawed cells were incubated 30 minutes at +4c with antibodies and with Live/dead Fixable Green Dead Cell Stain (at dilution of 1:500; ThermoFisher) that were diluted in in Brilliant Stain Buffer (BD Bicoscience). When applicable the cells were incubated with MR-1 tetramer (1:2500) in +37c for 45 minutes and washed before continuing to other surface markers. For staining of transcription factors and Ki67 the cells were permeabilized after surface staining with FoxP3 transcription factor staining set (eBioscience) for and with Fixation/Permeabilisation Solution kit (BD Bioscience) for detection of intracellular cytokines as instructed by manufacturer. The samples were run using LSR Fortessa (BD Biosciences) and analyzed with FlowJo (BD Biosciences, LLC). The antibodies used in the study are shown in Supplementary Table X. Optimal concentration for antibodies was titrated with live PBMCs.

### Cell separation

Purification of CD3^+^ T cells or CD8^+^ T - cells from freshly isolated PBMCs was done with Pan T cell or CD8 Microbead isolation kit using LS Columns (both Miltenyi Biotec). Purity of CD3^+^ cells was on average 97 (+/-1,2)% purity and CD8s 93 (+/-5,1)%. CD4^+^CD25^+^CD127^−^ cells were sorted from freshly isolated PBMC with BD FACSAria II instrument (BD Biosciences). Prior sorting CD4 cells were enriched with Human CD4^+^ T Cell Enrichment Cocktail (Stemcell Technologies) when isolating CD4^+^CD25^+^CD127^−^ for suppression analysis.

### IL-2 induced STAT5 phosphorylation

Intracellular staining for phosphorylated STAT5 (pSTAT5) was performed on thawed PBMCs allowed to recover for 16 hours in complete medium. Cells were counted and seeded at 150,000 cells per well in 96-well plates at a volume of 100 μL in complete medium. After 30 minutes of incubation at 37°C, cells were stimulated with IL-2 (200 U/mL for 7.5, 15 or 30 minutes and fixed for 10 minutes at 37°C with prewarmed paraformaldehyde (at a final concentration of 2%). Cells were then washed twice and permeabilized with prechilled (−20°C) BD Phosflow Perm Buffer III (BD Biosciences). After 30 minutes on ice, cells were washed twice with 2% FBS in 1xPBS and stained for 35 minutes on ice with antibodies Cells were washed twice with 2% FBS in 1xPBS and suspended in the same buffer for analysis.

### Evaluation of Treg suppressor capacity

CD4^+^CD25^+^CD127^−^ Treg cells were incubated for 6 days with carboxyfluorescein diacetate succinimidyl ester–labeled autologous responder T cells in ratio of 1:1. Anti-CD3/anti-CD28 beads (Life Technologies) were used as stimulus. CD4^+^cells were analyzed and the suppression percentage was calculated with the following formula: 100 − ([% proliferation in presence of Treg/% proliferation in absence of Treg] × 100).

### T cell activation cultures

TCR activation for freshly isolated cells in vitro was done in flat-bottom plate coated with unconjugated CD3- and CD28-antibodies (Immunotools) and cells were cultured in CTL test media (Immunospot). After overnight stimulation Brefeldin A (BD Biosciences) was added for the last 6 hours of stimulation after which the cells were collected for intracellular cytokine staining. Similar stimulation without brefeldin was done for RNASeq. After 24h stimulation cells were collected for RNAseq and stored at −80c before analysis.

MAIT cell stimulation with *E. coli* and *Candida albicans* was done as previously described (*39*). Briefly, microbes were fixed with CellFIX (BD Biosciences) and added to the cell culture in concentration of 6 ×10^6^ CFU for *E. coli* and 3 ×10^6^ CFU for *C. albicans*. Unconjugated anti-CD28 antibody was added after one hour of culture with the fixed microbes. Cells were incubated for a total of 24 hrs. For a positive control, a commercial PMA/ionomycin preparation was used (Leukocyte Activation Cocktail, with BD GolgiPlug™, BD Biosciences) for six hours.

### NanoString analysis of patient PBMCs

RNA extraction was performed with RNeasy Mini Kit (Qiagen Hilden) as instructed by manufacturer from snap-frozen PBMCs, and 100 ng RNA in 5 µl volume was taken for NanoString gene expression analysis (NanoString Technologies). Our custom gene set consisted of 45 genes targeting IFN-regulated genes, JAK/STAT and NFkB signaling related genes. Also five housekepping genes were included in the analysis. Further details of the code set, hybridization, scanning and data analysis are described elsewhere(*25*).

### 3’ RNA-seq

RNA extraction was done with Qiagen miRNeasy Micro Kit (Qiagen Hilden). Quality and quantity of the extracted RNA samples was analyzed with 2100 Bioanalyzer using RNA 6000 Pico Kit (Agilent, Santa Clara, CA, USA). Single-indexed mRNA libraries were prepared with minimum input of each sample with QuantSeq 3’ mRNA-Seq Library Prep Kit FWD (Lexogen Gmbh) according to user guide version 015UG009V0240 or 015UG009V0220. For CD3+ and Treg samples samples, globin mRNAs were removed from samples with Globin Block Module (Lexogen) and 6 bp Unique Molecular Identifiers (UMI) introduced with UMI Second Strand Synthesis Module (Lexogen) for detection and removal of PCR duplicates. Quality of libraries was measured using 2100 Bioanalyzer DNA High Sensitivity Kit (Agilent). Sequencing was performed with HiSeq 2500 System (Illumina) in high output run mode using v4 chemistry. Read length for the paired-end run was 2×101 bp and target coverage of 5 M reads for each library. QuantSeq 3’ mRNA-Seq Integrated Data Analysis Pipeline on Bluebee® (Lexogen Gmbh) was used for preliminary quality evaluation of the RNA sequencing data and to obtain gene specific read counts. Data were analyzed utilizing Chipster v3.14 (54). Differential expression analysis was done using EdgeR for multivariate experiments either with 1 main effect (disease status in analysis of non-treated samples) or with two main effects (disease status and sample pairs in treated vs. untreated analysis) with TMM-normalization using Log2 transformation. Genes with <5 counts in <2 samples were removed from analyses. An adjusted p-value (FDR) of 0.05 was used as limit for significantly differentially expressed genes. Correlation and volcano plots were generated using the ggplot2 package in R. For pathway analysis data were analyzed through the use of IPA (QIAGEN Inc., https://www.qiagenbioinformatics.com/products/ingenuitypathway-analysis).

### Monocyte activation

Thawn human PBMCs were suspended in RPMI 1640 (Biowhittaker/Lonza) media, supplemented with 25 mM HEPES (Lonza), 100 U/ml penicillin and 100 µg/ml streptomycin, L-glutamine and 10% fetal calf serum (all from Gibco) and incubated o/n at +37°C. The cells were activated with 1 µg/ml LPS (Sigma) for 4 hours and thereafter the NLRP3 inflammasome was activated by using 5 mM neutralized ATP (Sigma) for 45 min.

### Cytokine and cell death measurements cell, and measurement of inflammasome components with RT-PCR

ELISA kit for il-1β (high sensitivity, ThermoFisher) was used as instructed by the manufacturer to quantify cytokine concentrations from serum. Serum calprotectin levels were measured with clinically validated test in HUSLAB (Helsinki University Hospital, Helsinki, Finland). The mature, cleaved form of IL-1β was measured from cell culture supernatants using Human IL-1β/IL-1F2 DuoSet ELISA (R&D Systems). Cell death was analyzed from the cell culture supernatants using lactate dehydrogenase detection kit (Roche Diagnostics).

After monocyte activation, total cellular RNA was purified using RNeasy Plus Mini kit (Qiagen), followed by cDNA synthesis with iScript kit (BioRad). Quantitative PCR was performed from 5 ng of cDNA per reaction using HOT FIREPol Evagreen qPCR SuperMix (Solis BioDyne) and LightCycler96 instrument (Roche). See Supplementary Table 5 for the primer sequences. Relative gene expression was calculated using the 2(-ΔΔCt) method using *RPLP0* and *CASC3* as the housekeeping genes.

### Histology

The immunohistochemistry stainings of lymph node biopsies were performed in HUSLAB Pathology Department (Helsinki University Hospital, Helsinki, Finland) with clinically validated antibodies and protocols.

### Auto-antibodies

Autoantibodies were measured as previously described with luciferase immunoprecipitation analysis (*32*).

## Data Availability

Individual genomic data cannot be made public according to the European union GDPR regulations and our ethical approval. Genomic data can be made available to researchers based on a reasonable request. For access to the individual level genomics data please contact the data access committee of Institute for Molecular Medicine Finland (if.iknisleh@cad-mmif).

## Acknowledgements

The MR1 tetramer technology was developed jointly by Dr. James McCluskey, Dr. Jamie Rossjohn, and Dr. David Fairlie, and the material was produced by the NIH Tetramer Core Facility as permitted to be distributed by the University of Melbourne. This study was supported by grants from Emil Aaltonen Foundation, Sigrid Juselius Foundation, Finnish Medical Foundation and Academy of Finland (grant 308913). The authors thank Tamas Bazsinka and Sini Miettinen for technical assistance. Hetemäki designed and performed experiments, analyzed data, and wrote the original draft of the manuscript with Kekäläinen Kaustio and Kinnunen designed and performed experiments, analyzed data, and contributed to writing and editing of the manuscript Heikkilä, Keskitalo, Miettinen, Sarkkinen, Glumoff, Andersson, Kettunen, Vanhanen, Nurmi, Dunkel, Schlums and Kisand designed and performed experiments, analyzed data, and contributed to editing of the manuscript Otava and Syrjänen acquired the samples and clinical data and contributed to editing of the manuscript Eklund, Mäyränpää, Arstila, Bryceson, Peterson, Saarela, Varjosalo designed experiments, analyzed data, and contributed to editing of the manuscript Kekäläinen coordinated the study, designed experiments, analyzed data, and wrote the original draft of the manuscript.

## Supplementary Materials

Supplementary Table 1: Whole exome sequencing and filtration of data

Supplementary Table 2: Protein-protein interactions of HELIOS identified in biotin proximity ligation and ClueGo clustering of the results

Supplementary Table 3: List of differentially expressed genes in different RNAseq conditions

Supplementary Table 4: Pathway analysis of RNAseq data trough IPA

Supplementary Table 5: Supplementary list of reagents

## Supplementary Clinical Data

Patient 1 is a woman that was admitted to immunodeficiency clinic at her late 30s. He had suffered from recurrent *Candida albicans* vulvovaginitis with painful genital ulcerations for appr. 10 years before admission. The patient has had occasional aphthae also in her mouth and recurrent sinusitis. The diagnosis of common variable immunodeficiency was made and hypothyreosis diagnosis a year after that. The same year, some round, mildly enlarged lymph nodes were found in the right armpit and under the right pectoralis major muscle, histology showed cortical follicular hyperplasia.

Immunoglobulin replacement therapy was started two years after the diagnosis: with a weekly dose of 30‒40 ml the situation got significantly better and yeast infection relapses are now rare. After initiation of the immunoglobulin replacement therapy, she had a herpes zoster infection in the left chest area and she had two sinusitis with continuous sensation of nasal fullness. Weekly SCIG dose elevation to 60 ml did not help and antrostomy (FESS) on both sides was performed. While on SCIG, patient has been treated succesfully for *Helicobacter pylori*. Fecal parasites were examined due to persistent diarrhea and *Dientamoeba fragilis* was found and treated. Diarrhea persisted after succesfull eradication and a colonoscopy was made with normal macroscopic and histological findings.

Patient 2 is a relative of Patient 1 and he was examined at the immunodeficiency clinic first time at his 60s. During his working years, he had approximately two pneumonias yearly, after he has been examined for immunodeficiency he had one radiologically confirmed pneumonia two years ago. He has had a mild wound infection twice after surgery. Lichen planus of the upper extremities and gluteal area has reappeared over the years. At his 30s, Morbus Hodgkin was diagnosed in a lymph node of the left armpit. The lymphoma was treated with splenectomy and radiation therapy that resulted in hypothyreosis and mild fibrosis of the lungs, for which he uses intermittent glucocorticoid inhalations and gets occasional oral candidiasis. 16 years later, an enlarged lymph node was removed from the right inguinal area, histology showed lymphatic hyperplasia. *Helicobacter pylori* has been cleared with antibiotics. The patient has always had loose stools several times a day, colonoscopy two years after diagnosis showed mild irritation of the ileum but histology was normal.

| Clinical findings | patient 1 | patient 2 |
| --- | --- | --- |
| Viral infections |  |  |
| Aphtae | x | x |
| HSV | ~ | ~ |
| VZV (zoster) | x | I |
| HPV high risk | ~ | (-) |
| CMV | ~ | I |
| Bacterial infections |  |  |
| Recurrent sinusitis | x | ~ |
| Recurrent pneumonia | ~ | x |
| Surgical wound infection | ~ | x |
| HePy | x | x |
| Yeast infections |  |  |
| Mouth | ~ | x |
| Candida vaginitis | x | (-) |
| Parasitic infections |  |  |
| Dientamoeba fragilis | x | ~ |
| Giardia | ~ | ~ |
| Autoimmunity |  |  |
| Vitiligo | x | ~ |
| Lichen | ~ | x |
| Colitis | ~ | x |
| Hypothyreosis | x | x |
| Lymphatic tissue |  |  |
| Follicular hyperplasia | x | x |
| Lymphoma | ~ | x |
| Splenectomy | ~ | x |
| Vaccine responses |  |  |
| StPn polysaccharide | 4/10 | N |
| StPn conjugate | 2/9 | N |
| Menigococcus | (-) | N |
| Tetanus | low | N |
| Diphtheria | low | N |
| Measles | low | I |
| Rubella | N | I |
| Parotitis | N | I |
| Hib | (-) | N |
| HBV | low | (-) |
| HAV | N | (-) |
| Other |  |  |
| C3 | 1.47 low | 1.36 / N |
| Monosytosis | N | 0.81-0.92/12% |
| Interferon1 autoAb | N | elevated |
| Plasma IL2R | 535 N | 201 N |

HSV - Herpes simplex virus, S- HSVAb neg and no symptoms
VZV - Varicella zoster virus, zoster symptoms and VZV-IgG pos (patient 1), no symptoms and VZV-IgG pos (patient 2)
HPV - Human papilloma virus, high risk subgroups, PCR from cervix
CMV - P-CMVN<sub>h</sub> neg Patient 1
HePy - Helicobacter pylori, fecal antigens
StPn - Streptococcus pneumoniae
Hib - Hemophilus influenzae
HBV - Hepatitis B virus
HAV - Hepatitis A virus

---

**Supplementary Figure 1.**
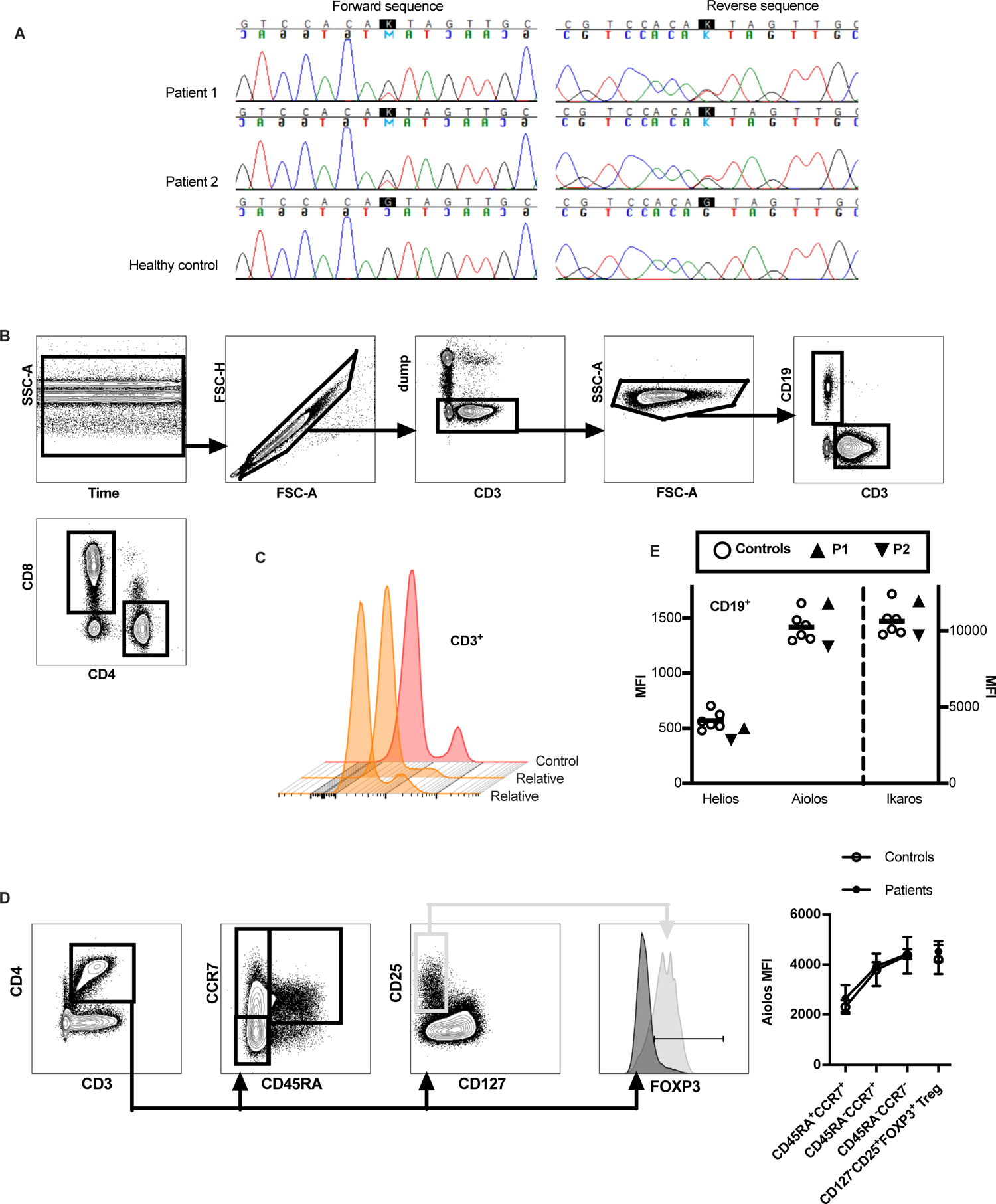
(A) Targeted capillary sequencing of cDNA derived from the RNA of patients and a healthy control, respectively. (B) Gating strategy to identify B cells, CD4+ T cells and CD8+ T cells from peripheral blood to calculate mean fluorescence intensity for transcription factors HELIOS, AIOLOS and IKAROS. CD19+CD3-B cells and CD19-CD3+ T cells were gated from live CD14-lymphocytes. CD4+CD8- and CD8+CD4-T cells were further identified from the T cell population. (C) Histogram showing HELIOS expression in two healthy relatives of patients and representative unrelated healthy control. (D) Gating strategy and mean AIOLOS MFI in patients compared to healthy controls (n=6) when examining AIOLOS expression in different subpopulations of CD4+ cells. Gating strategy is shown for a healthy male control (A, D). (E) Mean fluorescence intensity (MFI) for transcription factors in CD19+ B cells. Expression in CD4+ and CD8 + T cells is shown Fig 1C. P1=patient 1, P2=patient 2, (black triangles). Dump = dead cell marked, CD14.

**Supplementary Figure 2.**
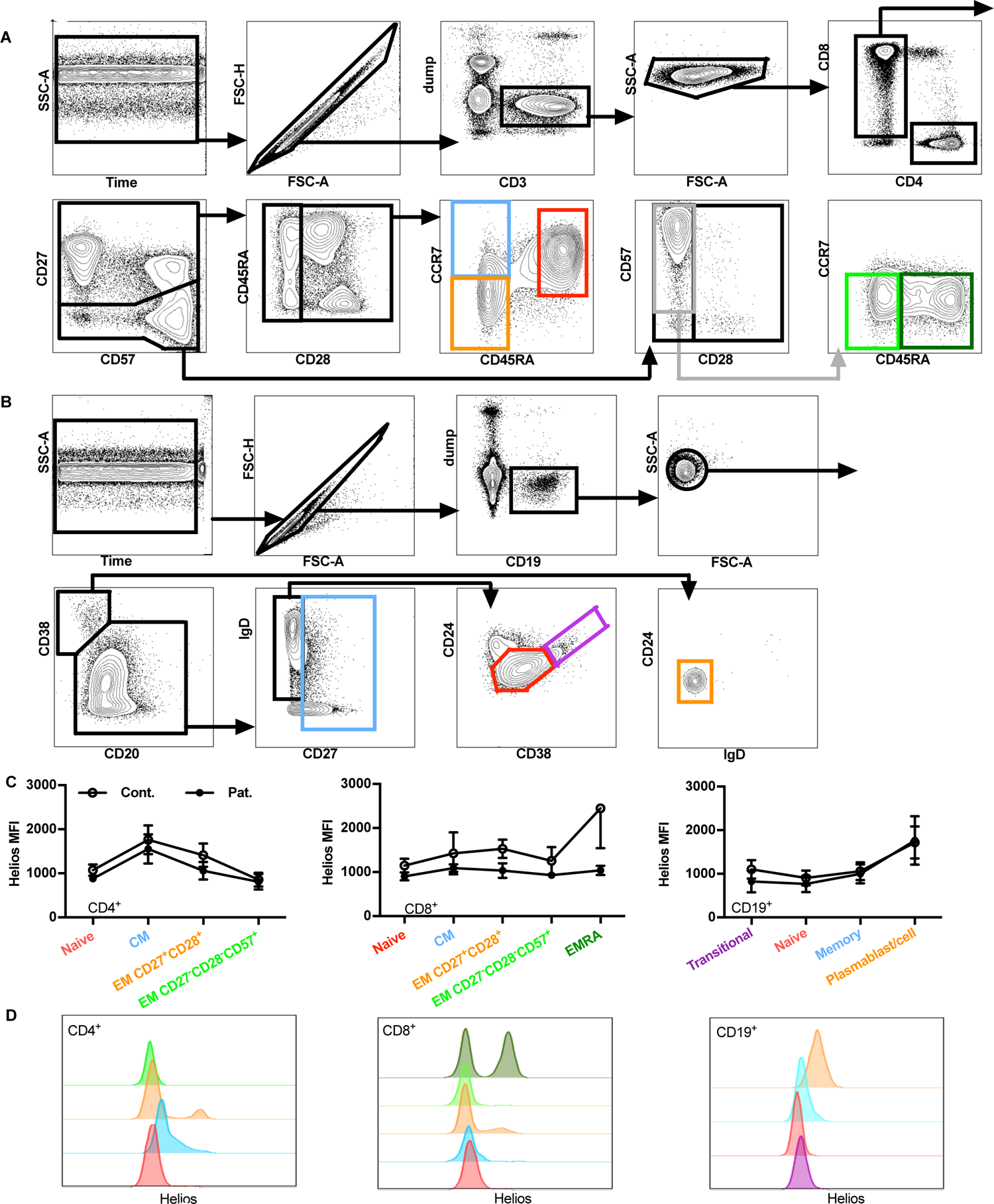
(A) Gating strategy to identify T and B cell maturation phases from peripheral blood and to measure their HELIOS expression. CD4+CD8- and CD8+CD4-T cells were gated from live CD14-CD19-CD3+ cells. These populations were divided by the expression of CD27 and CD28. CD27+CD28+CCR7+CD45RAhi naive (red) CD27+CD28+CCR7+CD45RA-central memory (blue), CD27+CD28+CCR7-CD45RA-effector memory (orange), CD27-CD28-CCR7-CD45RA-CD57+ effector memory (light green) and CD27-CD28-CCR7-CD45RA+CD57+ effector memory RA+ (dark green) cells were identified. Gating strategy in CD8+ cells is shown for a healthy male control. This gating strategy was also utilized to identify T cell populations depicted in Fig 2A&B. Dump = dead cell marked, CD19, CD14. (B) Cells of B cell lineage were identified as live CD3-CD14-CD19+cells. Transitional B cells were defined CD20+CD27-IgD+CD38hiCD24hi (violet), naive B cells CD20+CD27-IgD+CD38medCD24med (red) and memory B cells CD20+CD27-(blue). Plasma cells and -blasts were CD20-Cd38hiCD24-IgD-(orange). Gating strategy is shown for a healthy female control. Dump = dead cell marked, CD14, CD3. (C) Mean fluorescence intensity of HELIOS in respective lymphocyte populations in healthy controls (n=6) and patients. (D) Histograms depicting HELIOS expression in respective lymphocyte populations in a representative healthy control.

**Supplementary Figure 3.**
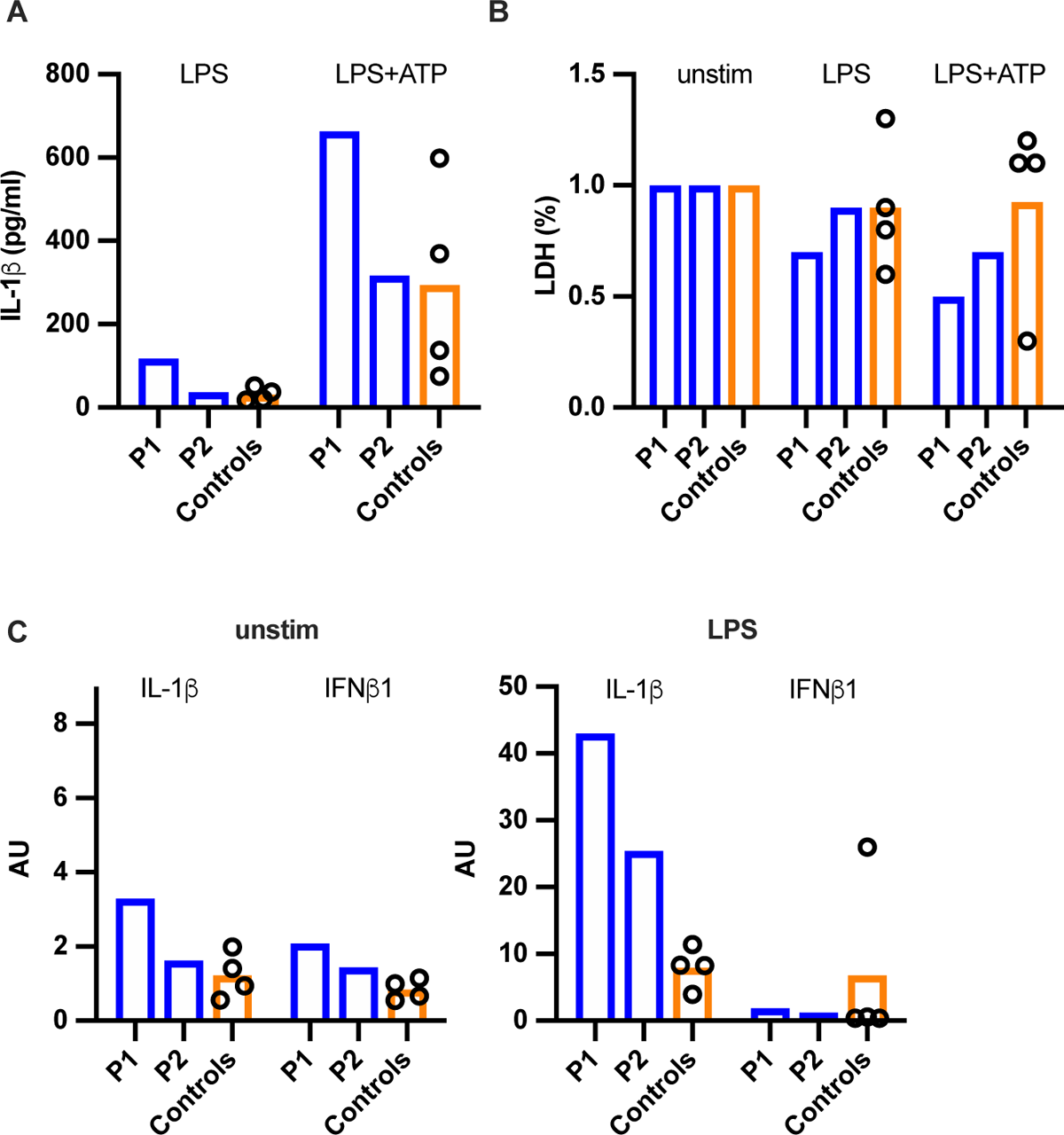
(A) PBMC of patients and healthy controls (n=4) were stimulated with LPS or combination of LPS and ATP, and il-1β and (C) lactate dehydrogenase (LDH) were measured from culture supernatants. (D) Expression of il-1β and IFNb1 were measured by quantitative PCR from PBMC without or with LPS stimulation. P1=patient 1, P2=patient 2.

**Supplementary Figure 4.**
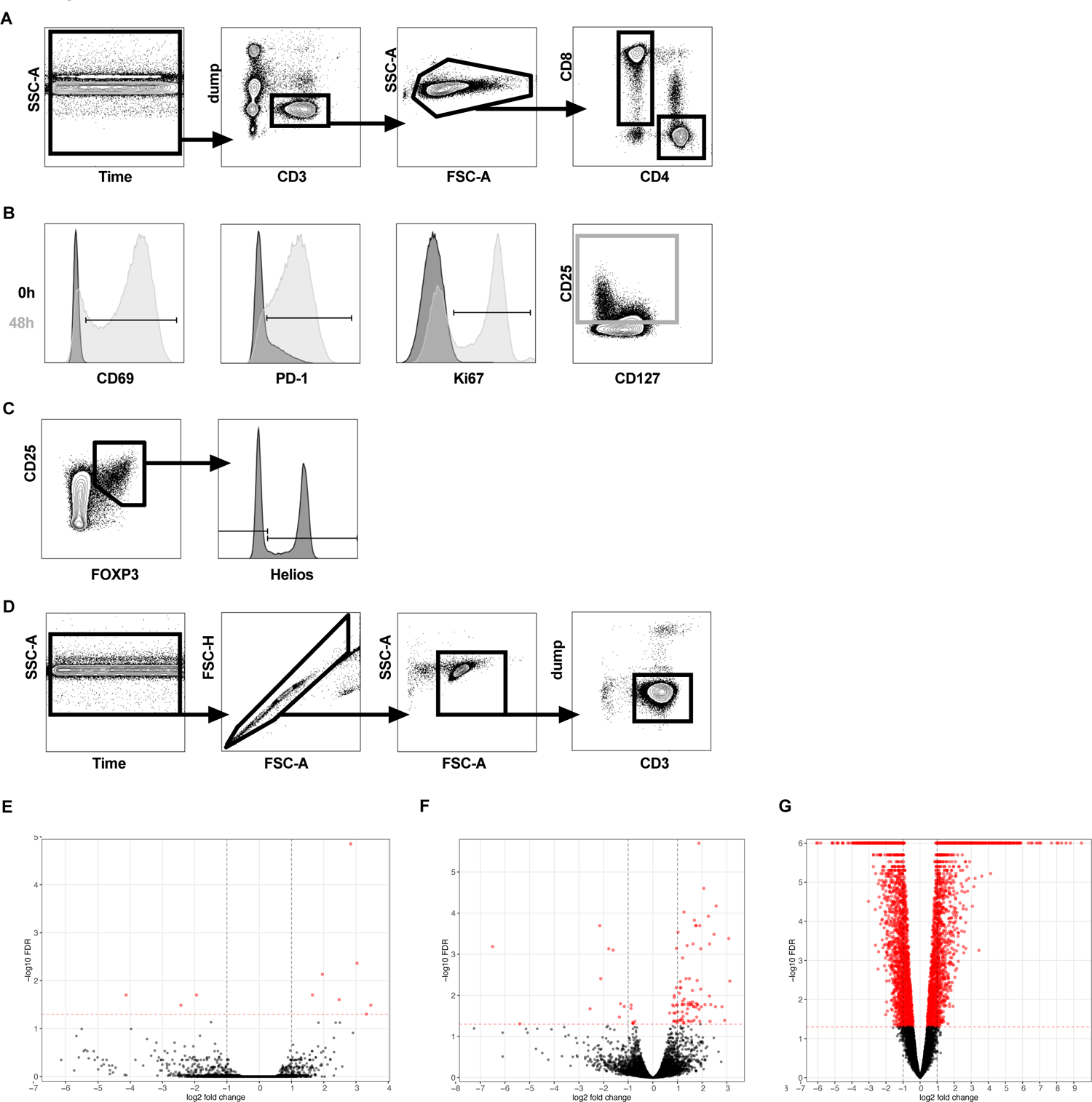
(A) Gating strategy to evaluate changes in activation markers and in cytokine production after stimulation with anti-CD3 and anti-CD28 antibodies. From live CD19-CD14-CD3+ lymphocytes with CD4+CD8- and CD4-CD8+ T cells were identified. Doublet discrimination was not performed as this would have resulted to unwanted exclusion of stimulated dividing cells. (B) Expression of CD69, PD-1 and Ki67 was measured in different time points, here expression in CD4+CD8-cells ex vivo (black) and after 48h of stimulation (light gray) is shown. (C) In different flow cytometry panel CD4+CD8-cells were identified with identical gating strategy as in A and CD25hiFOXP3hiHELIOS- or HELIOS+ cells were gated. IL-2 and IFN expression was then measured as shown in Fig 3D. (D). Additionally, the purity of CD3+ T cells was evaluated after bead purification. Gating strategy is shown with samples from a healthy female control (A-D). (E) Volcano plot of differentially expressed genes (red dots) in patients compared to healthy controls (n=5) in 3’RNA-seq data from purified CD3+ T cells without or (F) with anti-CD3-CD28 stimulation. (G) Volcano plot showing differentially expressed genes in anti-CD3-CD28 stimulated cells compared to unstimulated cells. Dump = dead cell marked, CD19, CD14.

**Supplementary Figure 5.**
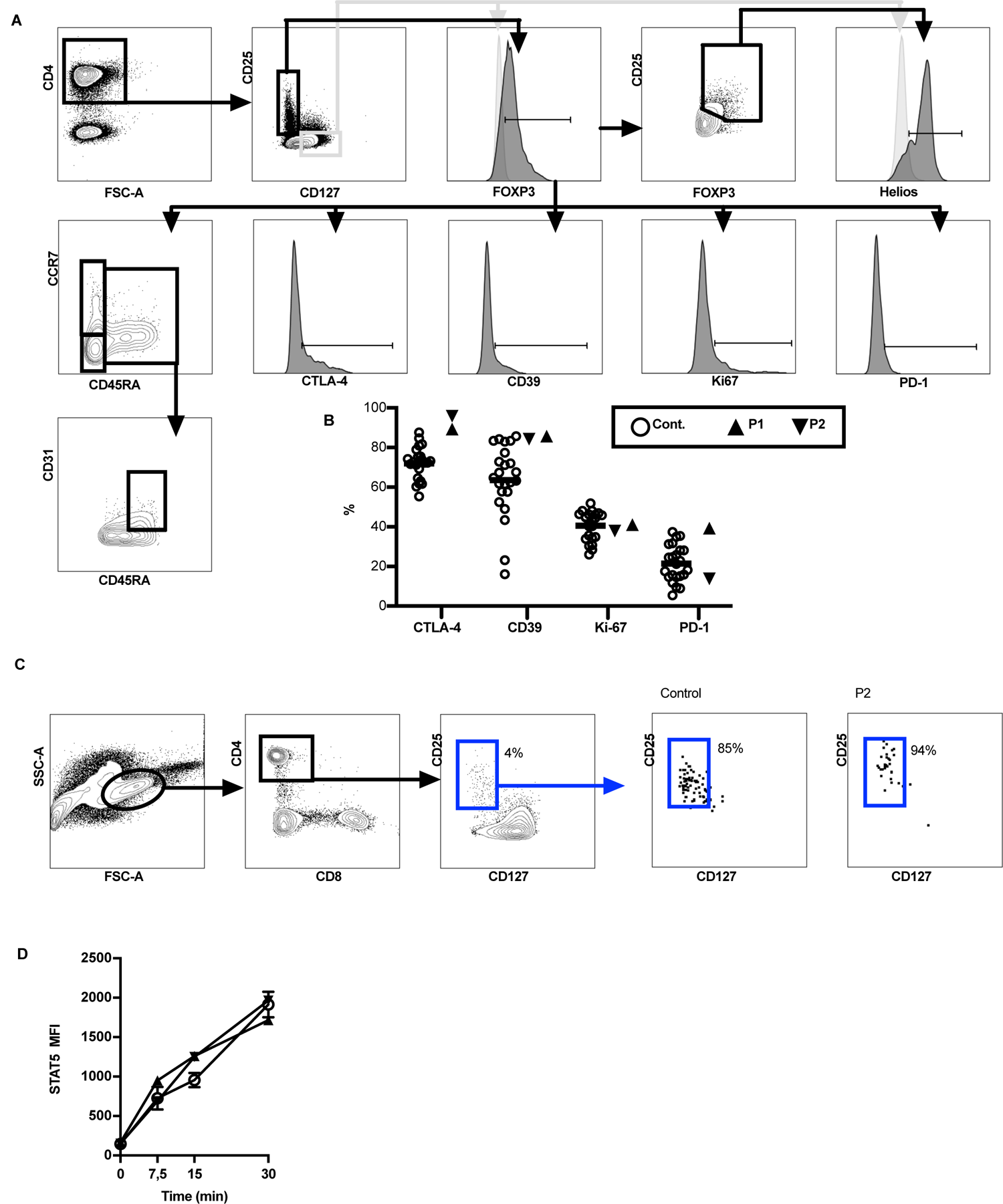
(A) Gating strategy to identify Tregs from peripheral blood. Live CD14-CD19-CD3+ T lymphocytes were gated as in Suppl Fig 2A. CD4+CD25+CD127-FOXP3+ Tregs were identified and the proportion of these cells expressing CTLA-4, CD39, Ki67 and PD-1 was measured. CD45RA+ cells shown in Fig 4B account for all the other CD45RA+ cells except CD45RAhiCD31+ cells and CCR7+CD45RA- and CCR7-CD45RA-were gated as shown above. Also more detailed analysis to identify CD25hiFoxP3hi Tregs and HELIOS+ cells from these cells was performed. Gating strategy is shown for a healthy male control. (B) Expression of different surface markers of CD4+CD127-CD25hiFOXP3hi Tregs in patients and controls. (C) Sorting strategy to CD4+CD127-CD25hi Tregs and purity in representative healthy control and patient 2 (P2). (D) STAT5 MFI in response to IL-2 stimulation at different time points in patients and healthy controls (n=3).

**Supplementary Figure 6.**
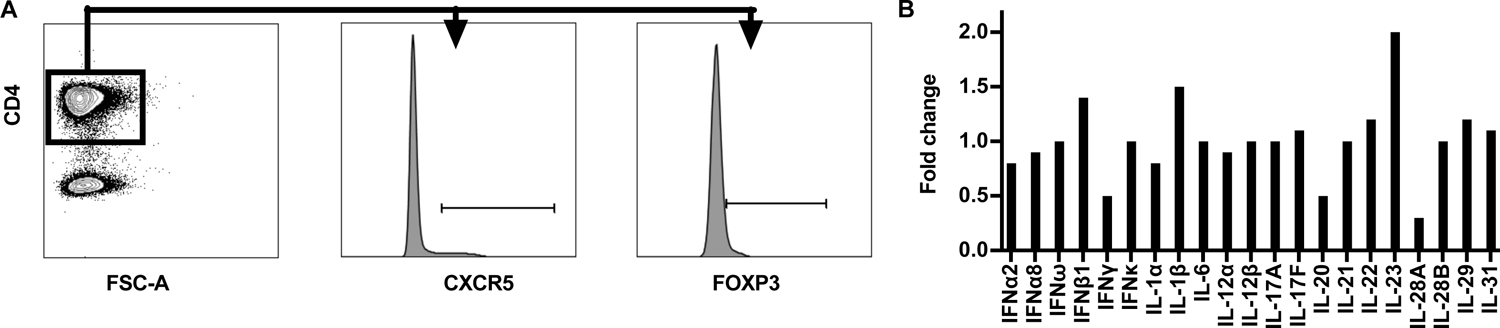
(A) Gating strategy to identify TfH cells from peripheral blood. Live CD14-CD19-CD3+ T lymphocytes were gated as in Suppl Fig 2A. CD4+ cells expressing CXCR5 and FOXP3 were identified from them. Gating strategy is shown for a healthy male control. (B) Auto-antibodies against cytokines in patient 1. Fold change compared to mean of healthy controls (n=5) is shown.

**Supplementary Figure 7.**
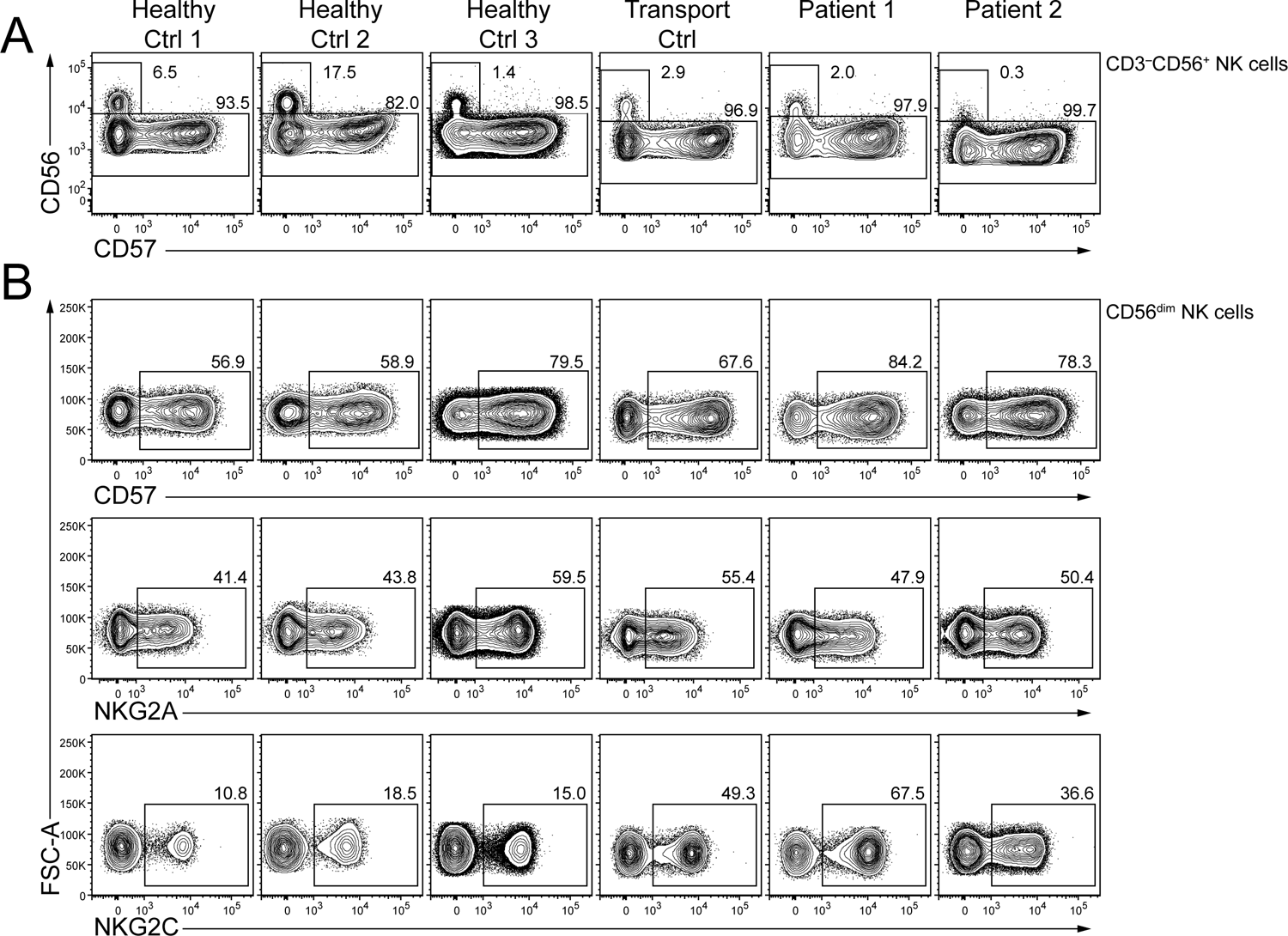
(A) Proportions of CD56brightCD16- and CD56dimCD16+ NK cells in controls and patients, (B) Proportions of fully mature CD57+, naïve NKG2A+, and adaptive-like NKG2C+ CD56dim NK cells in patients and controls.

**Suppl Fig 8.**
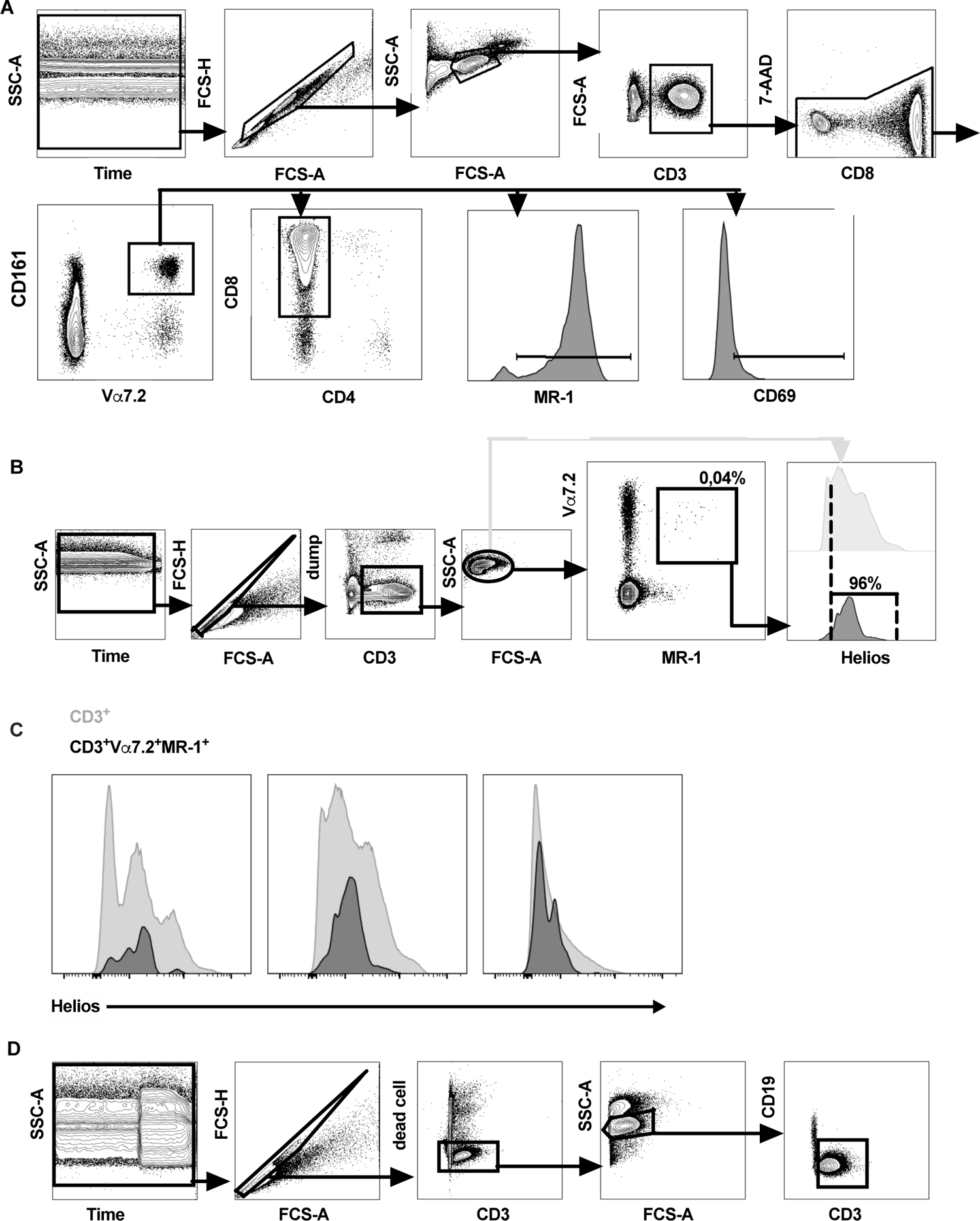
MAIT (A) Abundance and functional characteristics of MAITs were analyzed from freshly isolated PBMCs. From live CD3+Vα7.2+CD161+ MAITs expression of MR-1 and CD69 and the proportion of CD8+CD4-cells were evaluated. (B) Developing MAITs were identified from thymus as live CD19-CD14-CD3+Vα7.2+MR-1+ thymocytes. Cut-off for HELIOS positive cells was determined from HELIOS expression of all live CD3+ thymocytes (light gray). (C) Histograms showing HELIOS expression in all CD3+ thymocytes (light gray) and CD3+Vα7.2+MR-1+ thymocytes (black). (D) From duodenum and colon live CD19-CD3+ were first gated and MAITs identified as Vα7.2+CD161hi cells as shown in Fig 6 D&E. Gating strategy is shown with PBMC from a healthy female control (A) with a thymic sample from a male infant (B) and duodenal sample from a healthy female in her 30s (D). Dump = dead cell marked, CD19, CD14.

